# A demonstration of cone function plasticity after gene therapy in achromatopsia

**DOI:** 10.1101/2020.12.16.20246710

**Authors:** Mahtab Farahbakhsh, Elaine J. Anderson, Andy Rider, John A. Greenwood, Nashila Hirji, Serena Zaman, Pete R. Jones, D. Samuel Schwarzkopf, Geraint Rees, Michel Michaelides, Tessa M. Dekker

**Affiliations:** UCL Institute of Ophthalmology, University College London, UK; UCL Institute of Cognitive Neuroscience, University College London, UK; Experimental Psychology, University College London, UK; School of Ophthalmology, City University, London, UK; School of Optometry & Vision Science, University of Auckland, NZ; Moorfields Eye Hospital, London, UK; The Wellcome Centre for Human Neuroimaging, University College London, UK

## Abstract

Recent advances in regenerative therapy have placed the treatment of many previously incurable eye diseases within arms-reach (Ciulla et al., 2020). Achromatopsia (ACHM) is a severe monogenic heritable retinal disease that disrupts cone function from gestation, leaving patients with complete colour blindness, low acuity, photosensitivity, and nystagmus (Hirji, Aboshiha, et al., 2018). In non-primate animal models of ACHM, retinal gene-replacement therapy has successfully induced cone function in the young (Alexander et al., 2007; Carvalho et al., 2011), but it was yet to be determined if and when these therapies could effectively impact cone-mediated pathways in the human brain. Here we demonstrate in children with ACHM that gene therapy can yield substantial improvement in cone-mediated vision, via cascading effects on signal transmission from retina to cortex. To measure the effects of treatment in children with ACHM (CNGA3- and CNGB3-associated, all aged 10+ years), we developed novel visual stimuli, calibrated to selectively activate cone photoreceptors. We used these in behavioural psychophysics and functional MRI with population receptive field mapping, pre- and post-treatment. The results of treatment, contextualized against data from 12 untreated ACHM patients and 25 normal-sighted, revealed that six months post-therapy, two patients displayed novel responses to our cone-selective stimuli in the visual cortex, with a retinotopic organisation characteristic of normal-sighted individuals, not present in untreated ACHM. This was paired with significant improvement in cone-mediated perception specific to the treated eye, and self-reports of improved vision. Two other patients did not show a post-treatment effect, potentially reflecting individual differences in therapeutic outcome. Together, these data show that gene replacement therapy in humans with ACHM can activate dormant cone pathways despite long-term deprivation. This offers great promise for regenerative therapies, and their ability to trigger the neural plasticity needed to cure congenital vision loss in human patients.

## Main

Achromatopsia (ACHM) is a non-progressive recessively inherited retinal disorder in which disease-causing sequence variants in a single gene prevents cone photoreceptors from signalling. ACHM occurs in ∼1:30,000 births (Aboshiha et al., 2014; Johnson et al., 2004), with the most prevalent variants located in two genes, *CNGA3* (∼30% of European and US cases) and *CNGB3* (∼50% of cases) (Kohl et al., 2005). These genes encode the α and β subunits of the cone cyclic nucleotide-gated (CNG) channel respectively, both essential for the cone phototransduction cascade. As a result, vision in patients with ACHM is rod-dominated, and characterised by low acuity (6/36-6/60), insensitivity to chromatic contrasts, day-blindness, photophobia, and involuntary oscillation of the eyes (pendular nystagmus) (Hirji, Aboshiha, et al., 2018). Practically, patients have difficulty reading and recognising faces, do not perceive colour, and may wear sunglasses or darkened lenses to reduce discomfort from light exposure.

The retinal integrity of the two commonest forms of ACHM (mutations in *CNGA3* and *CNGB3*) have been studied in great detail (cross-sectionally and longitudinally), both with high-resolution optical coherence tomography to investigate retinal lamination and also cellular imaging to directly probe the photoreceptor mosaic *in vivo* (Dubis et al., 2014; Georgiou et al., 2019; Hirji, Aboshiha, et al., 2018; Hirji, Georgiou, et al., 2018). These studies have identified that although there is a marked reduction in cone cell density, all patients have residual cone cells that could be targeted for rescue, albeit with significant inter-subject variability in number.

ACHM is a promising candidate for genetic therapy, given its well understood genetic aetiology, availability of animal models, the presence of potentially viable cone cells, and the accessibility and low immune response of the retina to surgical intervention. The feasibility of using gene therapy safely to successfully treat inherited eye disease, was demonstrated recently with the first FDA and EMA approved gene therapy for RPE65-associated retinal dystrophy, Leber’s Congenital Amaurosis, a severe early-onset blinding disease (Ciulla et al., 2020). There are currently three phase I/II gene therapy trials for *CNGA3*-associated ACHM (NCT03758404, NCT02935517, and NCT02610582), and two phase I/II gene therapy trials for *CNGB3*-ACHM (NCT03001310 and NCT02599922).

Recently, the first published results of gene therapy clinical trials in 9 (NCT02610582) and, more recently, 2 (NCT02935517) treated adults with *CNGA3*-associated ACHM included various measures of visual acuity, photophobia, contrast sensitivity, flicker fusion, and colour thresholds after subretinal gene therapy (Fischer et al., 2020; McKyton et al. 2021). These studies showed modest improved function in the treated eye compared to the untreated eye for some measures and patients. Whilst this is promising and demonstrates treatment safety, small changes are hard to dissociate from confounding factors such as attention and task strategy-related effects, so it remains to be established if these truly reflect improved cone function after gene therapy. One reason for these modest effects may be that for the mature visual system, functional benefits of gene therapy may be limited by reduced retinocortical plasticity (Fischer et al., 2020; McKyton et al., 2021): In animal models of ACHM, gene therapy had substantially larger functional benefits when applied in young animals (Carvalho et al., 2011). In humans, studies on amblyopia demonstrate that detrimental effects of atypical visual experience on neural resource allocation and visual function become more entrenched with age (Holmes et al., 2011; Kiorpes, 2019). With life-long absence of cone function in ACHM, rods may appropriate neural resources normally reserved for cones (Baseler et al., 2002). For these reasons it is likely that gene therapeutic benefits in ACHM can be enhanced or unlocked by exploiting the inherent plasticity of the developing brain. Effectively testing this requires sensitive markers of gene therapy outcomes that can be used reliably with young patients, often with low visual acuity, nystagmus, and shorter attention spans – and these are currently lacking.

Here, we leverage a multi-modal approach that tests the neurofunctional impact of ocular gene therapy in childhood, linking changes in psychophysical estimates of cone sensitivity to functional Magnetic Resonance Imaging (fMRI) measures of cone signal processing in the developing visual cortex. This approach was developed to obtain large, reliable therapeutic effect sizes based on three key features. Firstly, the novel focus of this work on paediatric patients is important because treatment early in life enhances the scope for benefit. Secondly, by selectively activating cone photoreceptor-mediated pathways our stimuli induce qualitatively different responses in presence of cone function, so the measures are more specific to treatment targets than standard previously reported approaches that do not distinguish between rod and cone function. Thirdly, the complementary nature of our cone-selective psychophysics and fMRI tests, allows for quantifying and accounting for potential confounds of each method individually, offering a strong internal validity test. We employ this approach in children with ACHM (CNGA3- or CNGB3-associated) enrolled in phase 1/2 clinical trials investigating subretinal gene therapy with adeno-associated virus vectors expressing CNGA3 or CNGB3 (NCT03758404 and NCT03001310, see clinicaltrials.gov).

As data collection for this study and clinical trials is still ongoing, and trial outcomes are yet to be disclosed, we present measures on our tests in 4 paediatric patients of similar age (+10 years old) each tested before, and ∼6 months after receiving gene therapy (patient T1 and T2, with CNGB3 and CNGA3 variants respectively are reported in Main Text; patient T3 and T4 with CNGA3 variants replicating results of patient T1 and T2 are reported in Supplementary Materials 1). The treatment effects are contextualised against data from 11 untreated patients with ACHM, and 28 normally sighted control participants. Treated patients were selected for of this report based on their matching ages and data quality, and because they represent the range of outcomes observed with the multi-modal approach after treatment thus far.

## Results

We present data from four patients with AHCM who underwent a gene therapy currently under trial. Two patients (T2 and T4) demonstrated evidence of therapy-induced improvement in cone function 6 months after treatment. Before treatment, both patient’s psychophysical and functional brain imaging measures resembled those of other untreated achromatopsia patients (total n=12). Measurements after treatment, resembled those obtained from normal sighted controls (n=25), with a pattern unlikely to be explained by measurement confounds or false positives. Two other patients with ACHM of matched age undergoing gene therapy, demonstrated no such improvement, suggesting this multimodal approach can effectively track individual differences in outcome. Post-treatment results from patient T1 and T2 are reported in the Main Text. For corresponding results from patients T3 and T4, see Supplementary Materials 1.

### Behavioural Psychophysics

To test cone sensitivity psychophysically, we used a “silent substitution” approach to generate chromatic pairs with a range of cone photoreceptor contrasts, varying from high to low cone contrast. The red, green, and blue phosphor values of each chromatic pair were selected to increase or decrease the response of long (L) and middle (M) wavelength sensitive cone photoreceptors, whilst keeping rod photoreceptors silent (see Methods). Screen luminance was maintained within the cone-sensitive range throughout (0.6 to 13 cd/m^2^). Stimulus validation (See Supplementary Materials 2), however, revealed that the light spectra emitted by high contrast chromatic pairs induced some rod-contrast, likely due to measurement noise and imperfect correction for the screen’s non-canonical gamma function. In line with this, some higher contrast chromatic pairs were detected by ACHM patients, presumably using rod-based vision. This made it crucial to establish before treatment, which contrast levels were reliably below the detection threshold of patients with ACHM. Improved discrimination beyond the no-treatment baseline range, provides evidence for induced cone function after gene therapy.

We embedded chromatic pairs in a 4AFC target localisation task (Figure 1A&1C), as well as in a 2AFC movement discrimination task with the population receptive field (pRF) mapping stimuli (Figure 1B&1D). The latter test was included to ensure that psychophysics measurements were representative of the circumstances under which fMRI cone sensitivity measures were acquired. In both cases, a 1-up/1-down staircase was used (converging on 50% correct) to identify the lowest stimulus contrast that patients with ACHM could detect. Note that even the lowest cone contrast presented (Figure 1A) is above threshold and effortlessly visible to a normal-sighted individual, but invisible to those with untreated ACHM. Despite different chance levels and tasks, the two psychophysical measures showed good correspondence before treatment (Supplementary Materials 2 Figure S2b&S2c), suggesting they reliably captured the smallest noticeable rod photoreceptor contrast.

**Figure 1:**
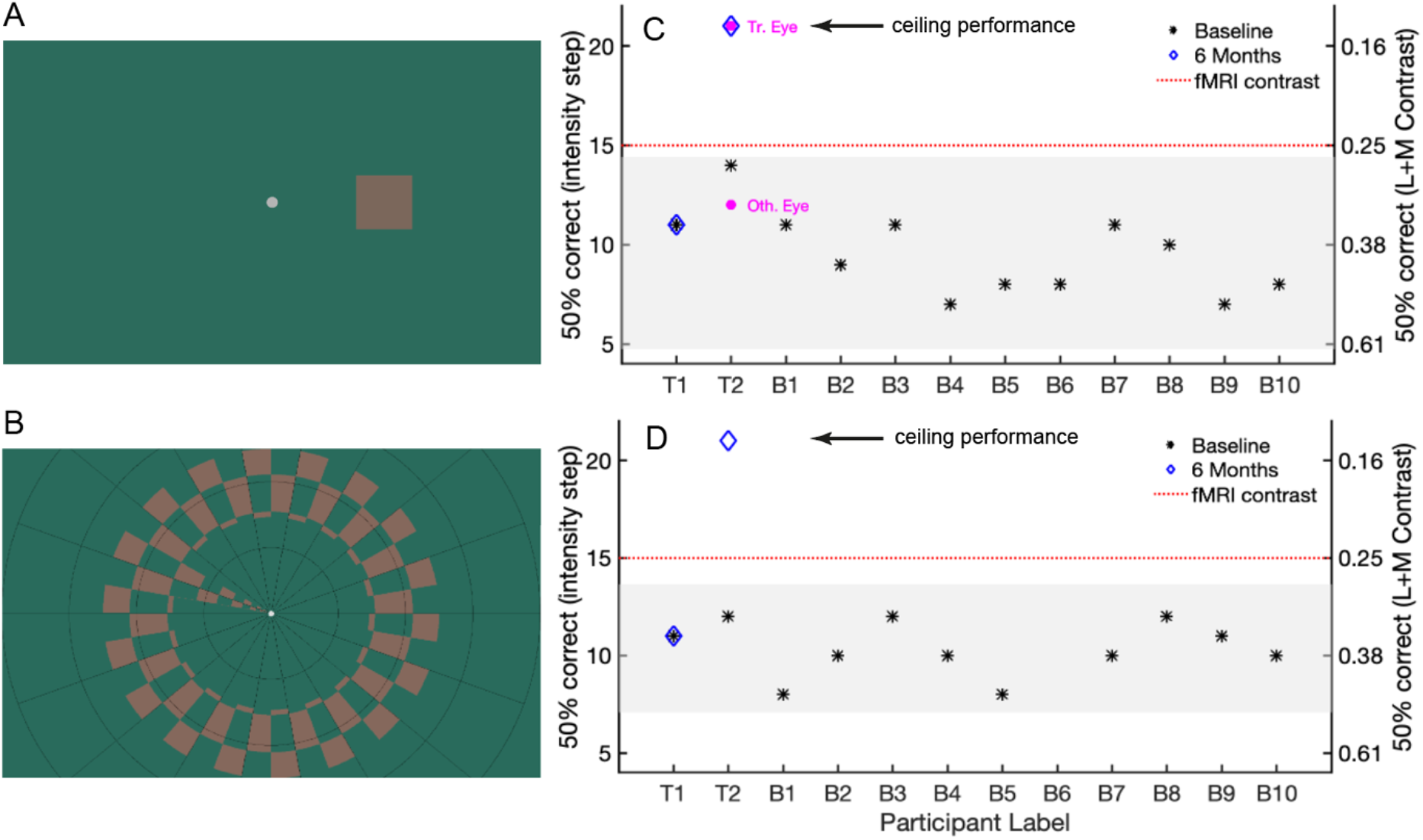
A) Psychophysics outside the scanner: example of the lowest cone-selective contrast tested psychophysically, embedded in 4AFC target localisation tasks. Participants judged the position of a target (size 3°), presented 6° to the left, right, above, or below centre, with unlimited time to search and uncontrolled gaze. Note that stimulus appearance is screen-dependent. B) pRF mapping stimulus presented inside the scanner: combined ring/wedge stimulus (max eccentricity 8.6°), depicted at the cone-selective contrast shown in the scanner (red dotted line, C&D). Participants detected target dimming events at fixation. After scanning, in a 2AFC psychophysics task, participants discriminated ring movement (inward/outward). Note that stimulus appearance is screen-dependent. C) Binocular 4AFC contrast discrimination thresholds (50% correct) for children with ACHM. Left y-axis indicates contrast detection thresholds in units of staircase step with decreasing stimulus intensity (1=highest contrast, 21=lowest contrast), with the right y-axis indicating the corresponding L+M cone Michelson contrasts (see Supplementary Material 2 for details). Stars indicate baseline measures for 12 untreated patients with ACHM. Shaded area: 95% prediction interval. Higher-contrast stimuli were above threshold for untreated patients with ACHM, likely due to imperfect rod silencing (Supplementary Materials 2 Figure S2a). Follow up measures ∼6 months after treatment, are shown for treated case study patients T1 and T2 (blue diamonds). 6-month measures of patient T2 were repeated monocularly for the treated and other, untreated eye (magenta circles). D) As in C but showing binocular contrast discrimination thresholds (at 50% correct) for the 2AFC task performed in the scanner.

Figure 1C & 1D present binocular baseline thresholds for 12 untreated patients with ACHM (black stars). For case study patients T1 and T2, we also present longitudinal measures collected ∼6 months after gene therapy (blue diamonds). Before treatment, both patients’ discrimination thresholds were in the range expected of children with ACHM. However, approximately 6 months after treatment, although the discrimination thresholds for patient T1 had remained unchanged, T2’s performance had improved to ceiling level - exceeding both their pre-treatment performance and baseline measures of 10 other untreated patients on both tasks (improvement exceeds 95% prediction interval for untreated patients, computed using the t-distribution quantile, shaded region in Fig 1C&D; see also Supplementary Materials 2 Figure S2b&c). Critically, this improvement was specific to the treated eye; T2’s monocular cone contrast discrimination was at ceiling for the eye that received gene therapy treatment, but remained at pre-treatment levels for the other eye (Figure 1C). In Supplementary Materials 1, we present psychophysics data from two additional patients tested after gene therapy (T3 and T4, respectively corresponding to B7 and B6 in Figure 1C&D). In patient T4 we observed a ceiling-level improvement in psychophysical performance in the treated eye as in patient T2, whilst in patient T3 discrimination threshold remained unchanged after treatment as in patient T1 (Figure S1a).

### Retinotopic Mapping fMRI

After quantifying individual differences in cone function psychophysically, we tested for concurrent evidence of cone-mediated signal processing in visual cortex. During fMRI, we presented participants in the scanner with one fixed cone contrast level (Figure 1B), embedded in a ring and wedge travelling checkerboard stimulus. Psychophysical measures and subjective reports confirmed that all 10 untreated patients with ACHM were unable to perceive this stimulus (Figures 1D, Supplementary Materials 2 Figures S2b & S2c). This confirms these stimuli were accurately calibrated to selectively stimulate cone photoreceptors whilst leaving the rods silent. We then used a population receptive field (pRF) mapping approach (Dumoulin & Wandell, 2008) to test for any cone-mediated responses in visual cortex (see methods).

We reasoned that the transmission of retinal responses to upstream visual cortex areas may be weak, if evoked for the first time in life following gene therapy. Therefore, to distinguish between non-retinotopically driven BOLD-signal fluctuations and retinotopically organised signal needed for functional vision, we compared the retinotopic structure of cone-mediated pRF maps with a rod-mediated retinotopic map from the same individual. To match rod-mediated maps as closely as possible across participants with and without functioning cones, we used stimuli designed to activate rods whilst keeping L- and M-cone photoreceptors silent. We refer to this map as ‘rod-mediated’, although S-cone signals may also contribute to these maps in control individuals (see Supplementary Materials 3). In normal visual development, polar angle tuning is spatially co-located for rod- and cone-mediated inputs in visual cortex, except around the foveal confluence, where rod-driven responses are lacking due to the absence of rod-photoreceptors in the retinal fovea (Barton & Brewer, 2015). For observers with healthy cone photoreceptors, we therefore expected high spatial correspondence between the cone- and rod-mediated polar angle maps. For untreated children with ACHM we expected no cone-mediated retinotopy, so there should be poor correspondence with the rod-mediated map, with emergence of well-aligned polar angle cone-mediated map structure after treatment providing evidence for new cone function. To maximise power to detect even weak cone-mediated signals in visual cortex, we did not apply a threshold to the pRF model goodness-of-fit when comparing cone- and rod-mediated maps (R^2^=0).

### Cone-mediated Retinotopic Map Structure

Patients with ACHM showed clearly visible retinotopic organisation in the rod-mediated (unthresholded) maps, as visualised for patient T1and T2 in Figure 2A. Their polar angle maps closely resembled those of the example control participant maps in overall layout. Note that map organisation is highly consistent within individuals. Before treatment, cone-selective pRF mapping evoked no visible retinotopic map in patient T1 and T2 (Figure 2B), as expected. After treatment, patient T1 still showed no discernible cone-mediated map, but patient T2 now demonstrated upper (red) and lower (green) visual field representations in expected cortex locations, aligned with the upper and lower fields in the rod-mediated map.

**Figure 2.**
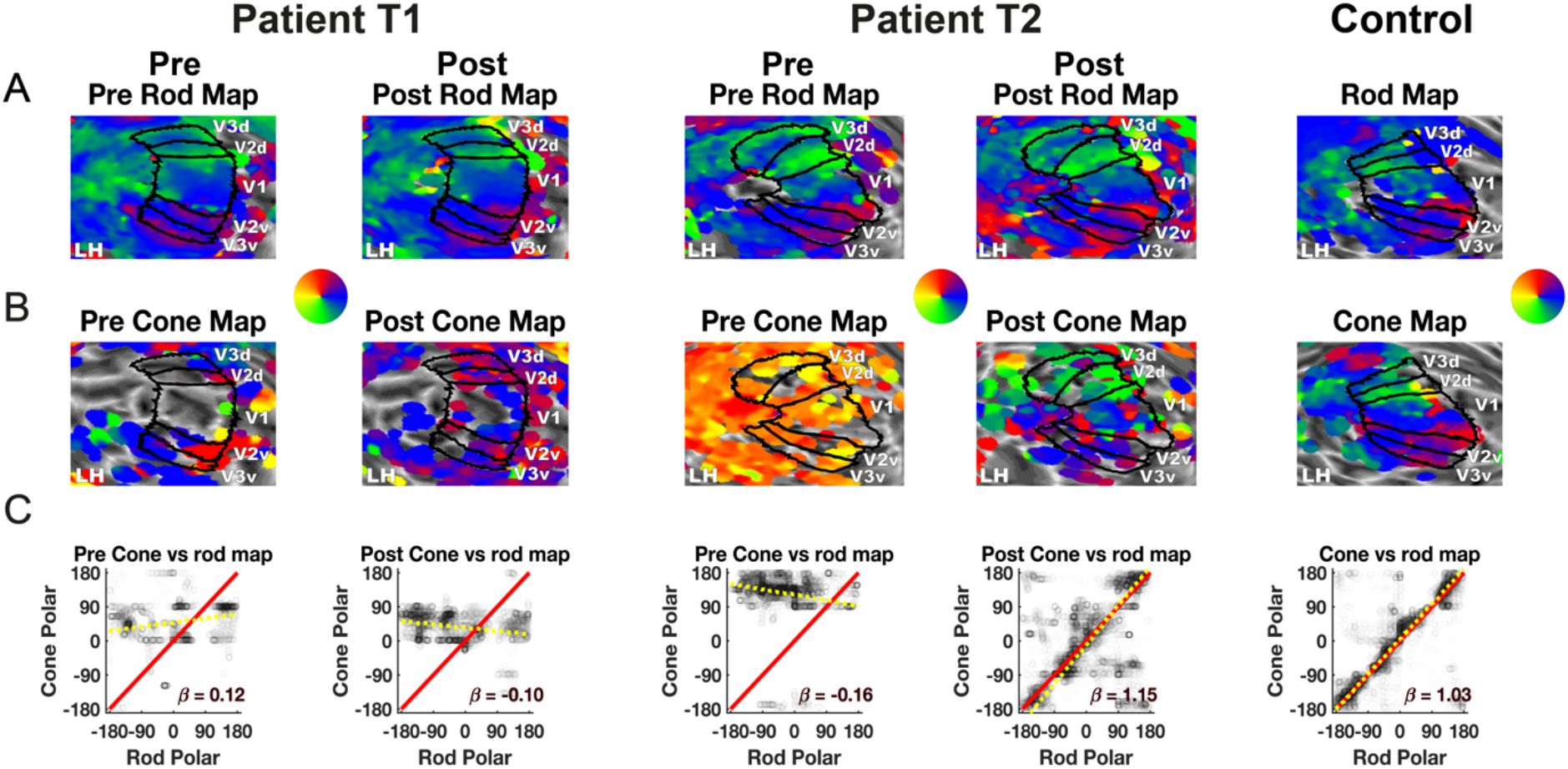
Unthresholded cone-mediated versus rod-mediated polar angle map organisation in areas V1, V2, and V3, before and after gene therapy in 2 patients, and an age-matched control participant. A) Rod-mediated polar angle estimates are projected onto the left hemisphere cortical surface, inflated to a sphere and zoomed in on. V1, V2, and V3 labels are drawn based on individual polar maps obtained in luminance contrast pRF scans. B) Cone-mediated polar angle map projected on the same hemisphere C) Rod-mediated polar angle values (x-axis) from the left and right V1, V2, and V3, scattered against cone-mediated polar angle values (y-axis). Red identity line indicates perfect correspondence between the rod and cone map. Yellow dotted line indicates the orthogonal linear regression fit, with slope β_orth_.

To quantify polar angle correspondence between maps, we plotted values from visual areas V1-V3 from the cone-mediated map against those from the rod-mediated map (Figure 2C). Close correspondence between maps in polar angle layout, is reflected in the clustering of data along the identity line. To test for presence versus absence of correspondence, we compared two orthogonal regression models, one following the identity line (slope β=1, intercept=0) and the other a horizontal line with a free parameter intercept to model relationship absence. To compute the strength of evidence for the correspondence model, we calculated the Akaike Weight (AIKW) (Burnham, & Anderson, 2002; Wagenmakers et al., 2004). For the control participant with normal cone function, there was strong evidence for correspondence across the cone and rod-mediated polar-angle maps (AICW_β=1_≈ 1), with clustering of data along the identity line (orthogonal regression slope *β*_*orth*,95CI_= [1.02:1.04]) and high circular correlation between maps (CorrCoef_Fisher-Lee_= 0.75, p<0.01; Pewsey et al., 2013) (Figure 2C, right). Before treatment, there was poor spatial correspondence between cone- and rod-mediated polar angle maps for both patients with ACHM (T1&T2 AICW_β=1_ ≈ 0), with low correlations (CorrCoef_Fisher-Lee_T1= −0.05, p<0.01, CorrCoef_Fisher-Lee_T2 = −0.12, p<0.01) and flat regression slopes (*β*_*orth*,95CI_T1 = [0.11:0.14]; *β*_*orth*,95CI_T2 = [-0.19:-0.14]). This dichotomy was replicated across 25 other normally sighted controls and 11 untreated patients (Supplementary Materials 4 Figure S4a). Note that the narrow range of cone-mediated polar angles in untreated patients, clustered around small eccentricities (see Figure 3), likely reflect subtle biases in the pRF fitting process uncovered by the low goodness-of-fit threshold (potentially related to the coarse-fit grid-search), rather than functional cone vision for these select field locations. Indeed, concurrent psychophysics data confirm that the cone-selective contrast used in pRF mapping was invisible to patients at all locations.

**Figure 3.**
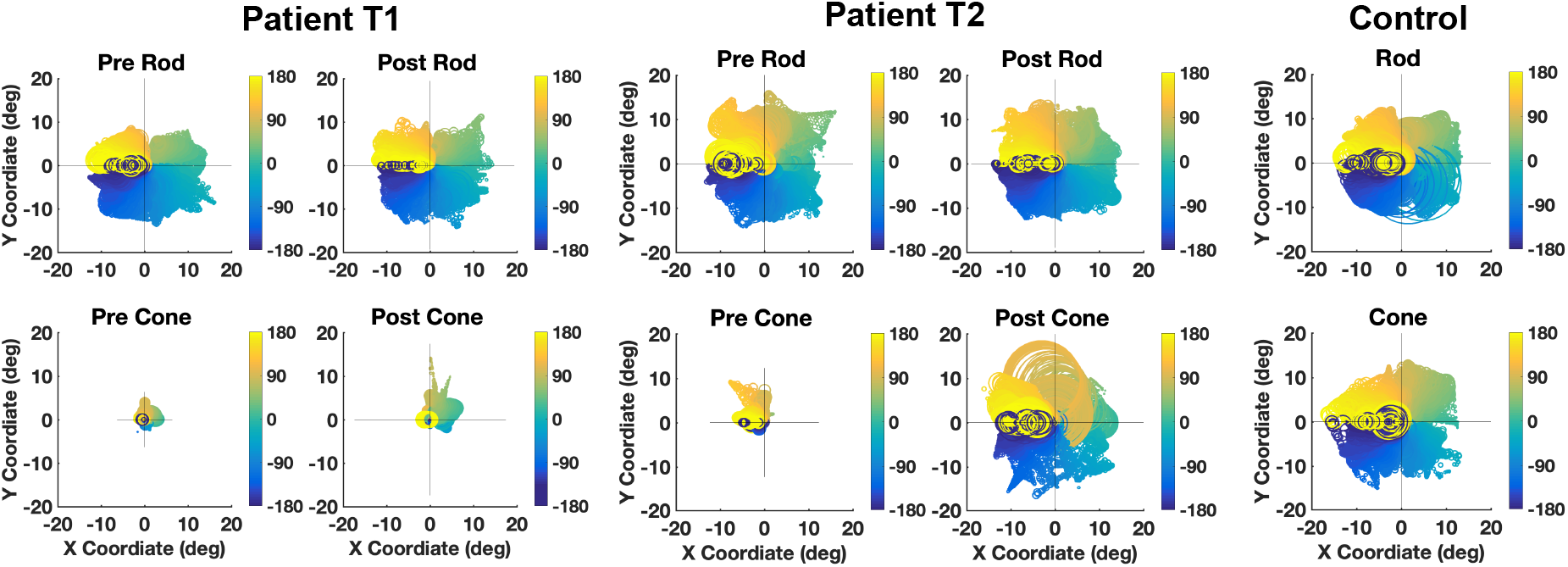
Visual field coverage before and after treatment for patient T1 and T2 and an age-matched control. Figures contain pRFs with unthresholded goodness of fit (R2 = 0) from V1, V2, and V3 extracted from the rod-mediated (top), and cone-mediated map (bottom) in left and right hemispheres. Colour indicates polar angle value. Circle size indicates receptive field size.

6 months after treatment, patient T1’s cone-mediated map still showed little correspondence with the retinotopic rod-mediated map (CorrCoef_Fisher-Lee_= −0.12, *β*_*orth*,95CI_= [-0.11:-0.9], AICW_*β*=1_ ≈ 0). In patient T2, however, a promising correspondence between these maps had emerged, albeit with a noisier relationship than in controls (CorrCoef_Fisher-Lee_= 0.3, p<0.01, *β*_*orth*,95CI_=[1.13:1.16], AICW_*β* =1_ ≈1). These results were replicated using the X and Y pRF position estimates that polar angle is computed from, so these results are not an artefact of polar angle circularity (Supplementary Materials 4 Figure S4b). While we also noted a substantial change in the cone-mediated eccentricity map of patient T2 after treatment, correspondence with the rod-mediated map was less clear than for the polar angle measure (Supplementary Materials 4 Figure S4c). It is possible that cone-mediated eccentricity map organisation may be more affected by life-long function loss or nystagmus. However, in patient T4 (Supplementary Materials 1), there was strong evidence for a new correspondence between the rod- and cone-mediated maps after treatment in both the polar angle (Figure 1Sb) and eccentricity measures (Figure 1Sc), showing it is possible to recover both visual field representation dimensions robustly after gene therapy. Like patient T1, patient T3 showed no evidence for emergence of cone-mediated map structure in visual cortex Figure 1Sb&c), in line with psychophysics measures of these patients, which also revealed no change.

### Cone-mediated visual field coverage

In Figure 3, pRFs measured from V1-V3 are plotted in visual space, with polar angle value indicated by colour, and pRF size indicated by circle size (R^2^=0). In all participants, pRFs derived from rod-mediated retinotopic maps tiled the visual field systematically, as did cone-mediated pRFs from the control participant. In contrast, cone-mediated visual field coverage in patients was poor before treatment. The small areas of near-foveal coverage likely arise from the same fitting artefacts discussed above rather than central cone vision in untreated ACHM (these disappear with a normal statistical threshold, e.g., goodness of fit at R^2^ >0.05). After treatment, visual field coverage had visibly increased in patient T2, but less so in patient T1. Population receptive field sizes derived from rod-mediated and cone-mediated mapping in patient T1 and T2 increased with eccentricity and were larger compared to those of control participants (see Supplementary Materials 5 Figure S5). Rod-mediated pRF sizes were reduced for both patient T1 and T2 after treatment. This is in line with recent findings of reduced pRF size in the treated and untreated eye of two adult patients with ACHM after gene therapy (McKyton et al., 2021). Smaller pRF size may indicate improved cortical acuity after gene therapy, but more data is needed to interpret this effect given the high test-retest variability of population receptive field size and its sensitivity to nystagmus (Clavagnier et al., 2015; van Dijk et al., 2016).

### fMRI Data Quality Control

It is important to ensure that longitudinal fMRI measures are not confounded by head or eye movements, as both requirements can be challenging for young individuals with nystagmus. For patients T1 and T2, head movement during scanning was minimal, with scan-to-scan movement remaining well below the 2.3mm^2^ voxel size pre- and post-treatment (Supplementary Materials 6 Figure S6). To assess fixation stability, we examined performance on the central fixation task, and variance of horizontal eye movements. We report horizontal eye movement because this is the dominant direction of these patients’ nystagmus, and because vertical eye movements were prone to blink-, eyelash, and scanner vibration artefacts (Supplementary Materials 7 Figure S7a). Task performance was high for both patients before and after treatment, as well as for the control participant (>95% in all pRF-mapping runs). Whilst horizontal eye movement variance in patients was about twice that of the control participant, both T1 and T2 were able to control their nystagmus well when fixation was required, with the largest median standard deviation across 1-second sections during fMRI runs 0.27 degrees, across all measurements (Supplementary Materials 7 Table S7, Supplementary Materials 7 Figure S7b). These control analyses suggest that fMRI data were well-matched in quality across participants, scanning conditions, and time points. Similar control analyses in patient T3 and T4 revealed that head- and gaze movement were also well-matched across these two patients although greater in both (Supplementary Materials Figure S1d, Figure S1e, Table S1).

## Discussion

Robust tests of visual function for children with inherited eye disease are essential for measuring the impact of new sight-rescuing therapies, particularly when treatment benefits are likely to be greater at young ages. Here we present a new multi-modal approach that leverages a combination of psychophysics and fMRI to address this need. We demonstrate the use of this approach for linking behavioural measures of cone function to cone-mediated signals in the visual cortex of children with genetically-confirmed ACHM, enrolled in gene therapy clinical trials targeting *CNGA3-* and *CNGB3*-associated ACHM.

Twelve patients with ACHM displayed no evidence of cone function across psychophysical and neural measures before the gene therapy intervention. They performed at chance when asked to detect chromatic-pair stimuli that selectively activate cone photoreceptors, and demonstrated no measurable cortical signal in response to these chromatic-pairs embedded in a pRF mapping stimulus. Cone-selective chromatic-pair stimuli that were invisible to all untreated patients with ACHM, were easily visible to normal-sighted control individuals. Together, this shows our study correctly targeted cone-selective information processing channels.

Approximately 6 months after gene therapy, the cone-photoreceptor targeting stimuli remained invisible to patient T1 but had become discriminable to patients T2. Selective to the treated eye, patient T2 improved to ceiling level on psychophysical tests of cone contrast sensitivity, indicating that performance on this task now matched that of individuals with normal cone function. In addition, there was strong evidence for cone-driven signals in visual cortical areas V1-V3 of patient T2, which had been absent prior to treatment. Activation patterns in response to the cone-selective stimulus also showed clear retinotopic organisation, now resembling maps from control children and adults with normal cone function. Given the highly organised nature of retinotopic responses in early visual cortex, it is most unlikely that these findings could have emerged from random noise fluctuations. Head- and eye-movements were measured throughout the pre- and post-treatment scan sessions and found to be comparable across conditions and time points. Therefore, the results observed in patients T2 after gene therapy, relative to patient T1, are highly unlikely to be driven by measurement confounds. Crucially, a replication of this pattern of results in patient T1 and T2 in two additional children with ACHM who underwent gene therapy (T3 and T4, Supplementary Materials 1), shows that these novel treatment-related effects can be replicated robustly in other cases, and do not reflect idiosyncratic characteristics of one specific individual.

From these early but striking results, we can conclude that gene therapy within the plastic period of visual development can successfully activate the dormant cone photoreceptor pathways in ACHM, and evoke visual signals not previously experienced by these patients. This shows that at least in some cases, a degree of neural infrastructure for useful cone function is preserved in ACHM after more than a decade of deprivation, well beyond the most sensitive periods for vision. These results are in line with studies showing that removal of bilateral congenital cataracts, lens-clouding that severely limits visual acuity, is most effective when done early in life, but that some visual recovery is possible even if treatment occurs late or in adulthood (Maurer, 2017; McKyton et al., 2015). In the case of ACHM, experience with rod-based vision may scaffold perceptual capacities that late recovered cone-mediated function may benefit from.

Whilst evidence for cone sensitivity clearly emerged after treatment in patient T2 and T4, cone-mediated fMRI measures remained less robust than those of controls. This may in part be because, by trial protocol, only the worse eye was treated, whilst our data was collected binocularly. Our measures in patients thus included potentially competing vision from the untreated and typically better eye. In addition, however, whilst the lowest cone contrast in the cone-selective stimulus set was immediately visible to controls with normal cone function, patient T2 and T4 took longer to detect the location of the stimulus before giving their response, even with the untreated eye closed. Thus, while aspects of cone function improved after treatment, the overall impact on broader visual function remains to be explored. Incidentally, patient T2 reported seeing “different” with their treated eye, mentioning perceived benefits for reading certain signs.

Our approach of measuring regenerative treatment outcomes across visual processing stages can provide important insight into the neural plasticity constraints beyond the retina itself that successful ocular gene therapies must overcome; Cone photoreceptor activation normally helps refine neural connections from retina to cortex through experience-dependent competitive processes (Huberman et al., 2008; Kiorpes, 2019). With cone activation missing from gestation in ACHM, enhanced competitive pressures from rod-dominated vision may alter the organization of visual pathways, and limit the scope for treatment efficacy. In line with this, one fMRI study reported that in three patients with ACHM, cortical areas that normally encode the foveal retina, which contains only cone receptors, were activated by more eccentric parts of the visual field that stimulate rod-containing retina (Baseler et al., 2002). This may indicate that rod-mediated signals can invade dormant cone-innervated cortex in ACHM. However, a similar shift in spatial tuning profile arises in normal sighted individuals from measurement artefacts at the edge of the rod scotoma (Barton & Brewer, 2015). It is therefore still unclear if reorganisation of cone signalling pathways occurs in ACHM, and if so, how and when. Because the success of gene therapy may depend in part on the degree to which post-retinal reorganisation is present and reversible, it will be important to investigate such processes in ACHM in the context of individual treatment outcomes.

A crucial question to address in future therefore, is what may explain individual differences in patient vision after gene therapy, how this changes across the lifespan, and how vision may be improved further by promoting neural plasticity. ACHM patients enrolled in gene therapy trials, including the four patients reported here, may differ on many variables, including retinal architecture, treatment dosage, genetic profile, and pre-existing retinal, post-retinal, and functional characteristics, some of which in turn vary with age. We anticipate that the fine-grained information obtained with fMRI combined with psychophysical measures of photoreceptor function, will be critical for elucidating how these genetic, neural, and developmental factors interact with regenerative therapies to permit visual recovery in patients with ACHM, and the many other congenital eye diseases for which new treatments are currently under development.

## Methods

### Participants

As the gene therapy trial and data collection are still ongoing, with treatment conditions and outcomes yet to be disclosed, we present interim measures from 2 paediatric patients who represent the range of results observed. The patients (patient T1 and T2, with CNGB3 and CNGA3 variants respectively), each were 10+ years old at the time of first visit (we keep exact ages undisclosed for confidentiality). T1 and T2 were tested twice, once before, and ∼6 months after treatment. Their data is presented in the context of MRI and psychophysics measures of 9 additional children with ACHM, including two other patients with post-treatment data (T3/B7 and T4/B6, see Supplementary Materials 1) and 2 adults (11 paediatric patients in total: Mean Age = 11.27, Range = 8-15 years, SD = 2.49, 2 adults in their twenties; all with genetically confirmed *CNGA3* or *CNGB3*-associated ACHM), and data from 28 normal-sighted controls (16 children: Mean Age = 10.62, Range = 6-14 years, SD = 2.60; 12 adults: Mean Age = 24.42, Range = 18-34 years, SD = 4.42). For one paediatric baseline AHCM patient, psychophysics data was not collected. For two normal-sighted participants the rod-mediated pRF map was not collected. Additional control normal sighted and ACHM patients were excluded from the analyses in case of excessive head movement (1 control, 2 patients), equipment problems (6 controls, 2 patients), or missing MRI measures (2 patients). All participants met MRI safety inclusion criteria and had no known neurological disorders besides, in patients, ACHM. Most paediatric ACHM patients in this study, were also enrolled in clinical trials NCT03758404 and NCT03001310. Informed consent was obtained from all parents and participants for taking part in this MRI study, and children themselves gave informed assent. Data collection for this MRI study had stand-alone ethics approval (separate from clinical trials) from the national ethics committee for patients (REC reference: 12/LO/1196; IRAS code: 106506) and the UCL ethics committee for normal sighted control participants (#4846/001).

### Apparatus

We used a Siemens Avanto 1.5T MRI scanner with a 30-channel coil (a 32-channel coil customised to remove view obstructions) to acquire structural and functional MRI data. Stimuli were presented on an MR-compatible LCD display (BOLDscreen 24, Cambridge Research Systems Ltd., UK; 51 × 32 cm; 1920 × 1200 pixels) viewed through a mirror in the scanner at 105 cm distance. Participants were lying supine in the scanner, with fixation stability recorded where possible via a mirror, with an Eyelink 1000 at the back of bore. Behavioural psychophysics was also performed in the scanner room, after scanning, under similar viewing circumstances as the functional MRI data collection. Hardware were controlled using custom MATLAB code (R2016b, MathWorks, Natick, MA, USA), via the Psychophysics Toolbox 3 (Kleiner et al., 2007). Data were collected binocularly in the scanner, to keep testing time feasible for young children. Whilst the treatment was applied monocularly, each eye projects to each cortical hemisphere. Improved cone photoreceptor signal transmission in the treated eye should therefore be measurable in affected retinal representations of bilateral visual cortex. Similarly, we expected functional benefits for the treated eye to be detectable with both eyes open, if potentially weakened due to ocular competition effects from the other eye.

### Cone-selective stimuli in psychophysics and functional MRI

To measure cone-mediated signal processing, we used the silent substitution approach to generate pairs of chromatic stimuli designed to selectively activate L- and M-cones whilst keeping rod activation constant (for details, see Estevez & Spekreijse, 1982; and Spitschan & Woelders, 2018). In brief, we computed transformation matrices to convert changes in the LCD screen’s red, green, and blue channel (RGB) voltages, into changes in L-cone, M-cone and rod photoreceptor stimulation, using measures of the screen’s RGB spectral outputs (made with a Spectrascan Spectroradiometer, PR-655, PhotoResearch Inc.) and the standard observer sensitivity functions for rod and cone photoreceptors (Stockman & Sharpe, 2000; Wyszecki & Stiles, 1982). This allowed us to calculate the change in RGB voltage required to independently increment or decrement L- and M-cone or rod photoreceptor activity by pre-specified proportions with respect to a baseline RGB value (mid-grey). We used this approach to generate chromatic-pairs that varied L- and M-cone photoreceptor activation whilst leaving rod photoreceptor activation constant. For these stimuli we required to silence only one type of photoreceptor (rods) rather than two, giving more freedom in the colour directions we chose. To account for any imperfect matching of rod activation (*e*.*g*., due to light measurement and correction errors, variations in rod sensitivity, or screen inhomogeneity), we kept the blue voltage constant (rods are relatively more sensitive to the blue channel than the L- or M-cones are) and only varied the R and G channels, thus shifting the stimulus variations towards longer wavelengths where rods are less sensitive and errors in rod equating are likely to be small. See Supplementary Materials 2 for validation measures. Neutral density filters were used to present these stimuli in the mesopic/low photopic light range (0.6-1.3 cd/m^2^ for psychophysics, 0.8 cd/m^2^ for fMRI), whilst keeping viewing comfortable for photosensitive patients.

### Rod-selective stimuli in functional MRI

Rod-selective stimuli, only used in the MRI scanner, were obtained using a similar approach. We used silent substitution to generate a chromatic pair that kept L and M contrast constant, but induced a contrasting response in rods. With 3 colour channels it is only possible to simultaneously silence two photoreceptor types, and the resulting stimulus was not controlled for S-cone contribution. Therefore, to reduce the S-cone response, we presented these stimuli at a very low light level (0.02 cd/m^2^) after dark-adaptation in the scanner. However, we cannot exclude the possibility that functioning cone photoreceptors may have been activated under these circumstances and contributed to the rod-mediated maps in individuals with S-cone function (for further details and validation measures, see Supplementary Materials 3).

### Behavioural Psychophysics

#### Each participant performed 2 psychophysical tests

##### 1) 4AFC localisation task

participants located a chromatic patch that induced a high L and M response (subtending 3° of visual angle) against a chromatic background that induced a low L and M response. The patch was presented at 6° eccentricity either left, right, above or below a central marker (0.4° VA) (Figure 1A). Participants were not required to fixate and had unlimited time to search. The cone contrast between target patch and background decreased gradually, following a 4-AFC 1-up/1-down staircase. Each adaptive staircase continued until at least 14 reversals had occurred (mean N trials = 27).

##### 2) AFC movement discrimination task

to assess the validity of these measures with the pRF stimulus, the cone contrast sensitivity threshold was also measured in a 2-AFC task with the cone-selective ring-and-wedge fMRI stimulus (Figure 1B). Participants detected movement of a checkerboard ring (inward/outward) or wedge (clockwise/anti-clockwise), reversing at 2Hz, in a 1-up/1-down staircase (converging at chancel level) until at least 8 reversals had occurred (mean N trials = 25).

### pRF mapping fMRI

Inside the scanner, participants first practised lying still whilst watching a cartoon that was paused whenever the researcher observed excessive head-movement via a face camera. Throughout each acquisition session, patients were constantly monitored for any signs of discomfort or movement via various cameras and a 2-way intercom. pRF mapping stimuli comprised of a simultaneous rotating ring and contracting/expanding wedge, each made up of cone- or rod-selective chromatic-pairs, embedded within a contrast-reversing checkerboard with 2Hz reversal rate (Figure 1B). Per run, the ring expanded/contracted for 6 cycles (48 secs/cycle) with logarithmic eccentricity scaling (van Dijk et al., 2016). The wedge (20° VA angle) rotated clockwise/anticlockwise for 8 cycles (36 sec/cycle). 20-second fixation baselines were embedded at the start, at the mid-point, and end of the run (total run duration 348 secs). The stimuli covered a maximum eccentricity of 8.6°, and moved to a new position each 1-second TR. The stimulus was overlaid with a small white central fixation dot (0.2° VA radius) and a black radial grid to encourage stable fixation. Participants completed 6 pRF mapping runs in total: 2 with cone-selective stimuli, 2 with rod-selective stimuli, and 2 with non-selective stimuli (*i*.*e*., standard luminance contrast-reversing checkerboard) used for delineation of cortical visual areas. High-resolution structural scans were obtained during a 15-minute dark adaptation phase, during which participants listened to an audiobook. The order of runs was pre-set and identical for each participant: 2 rod-selective, 2 cone-selective, 2 non-selective (luminance contrast) runs.

During the pRF mapping scans, participants indicated by button press when they detected a fixation target change from white to black. This task was presented as a rewarded “kitten rescue mission game” to promote engagement. Built-in calibration of the eye-tracker to quantify fixation stability, was not possible due to nystagmus. We therefore calibrated the camera in advance on a healthy eye, and for every other run we asked the patients to fixate on a 5-point custom calibration, used to calibrate eye-gaze measures post-hoc (see supplementary materials 7).

### MRI sequences

Functional T2*-weighted multiband 2D echo-planar images (Setsompop et al., 2012) were collected using a multi-band 28 sequence (TR = 1 ms, TE = 55 ms, slices = 348, flip angle = 75°, acceleration = 4) with a resolution of 2.3 mm isotropic voxels. A high-resolution structural scan was acquired using a T1-weighted 3D MPRAGE (1 mm^3^ voxel size, Bandwidth = 190 Hz/pix, 176 partitions, partition TR = 2730, TR = 8.4ms, TE = 3.57, effective T1 = 1000 ms, flip angle = 7 degrees). A lower-resolution MPRAGE was also obtained in the same orientation as the multiband sequence to aid co-registration between functional and structural images.

### MRI Data analysis

All functional data were pre-processed using SPM12 (http://www.fil.ion.ucl.ac.uk/spm). Functional volumes were realigned to the first image of each run to correct for head movement. All functional scans (collected pre- and post-treatment) were then aligned to the high-resolution structural scan collected pre-treatment. For accuracy, a low-resolution structural image with the same orientation as the functional volumes was used as an intermediate step to compute the co-registration matrix. FreeSurfer software (v5.3.0 with XQuartz v2.7.8,https://surfer.nmr.mgh.harvard.edu/) was used to construct 3D surface meshes for the right and left cortical hemispheres (Fischl et al., 1999), using the recon-all pipeline. Any holes or edges were corrected manually using FreeSurfer Freeview tool. Pre-processed functional data were projected onto the surface using the MATLAB toolbox ‘SamSrf’ v5.84 (https://osf.io/2rgsm/) for further analyses.

To model population receptive fields, we used a symmetric bivariate Gaussian model, with mean (x,y) representing the preferred retinotopic location, and standard deviation (σ) representing pRF size. To identify the pRF model (x, y, σ) that best predicts the measured time series, a two-stage fitting procedure was employed. In a coarse fitting step, data was smoothed along the cortical surface (Gaussian kernel fwhm = 5 mm), and a grid-search approach was used to identify model parameters that maximise the Pearson correlation between observed data and the pRF model’s predicted time course. Vertices with a good fit (R^2^ > 0.05) were entered as starting value in a fine-fitting step, in which Matlab’s fminsearch function was used to identify parameters that minimised the squared residual deviations between the model and unsmoothed data. Finally, best-fitting parameters were smoothed along the surface (fwhm = 3 mm), and X and Y position estimates were converted to eccentricity (distance from fixation) and polar angle.

### Regions of Interest

Visual regions were delineated manually by displaying the polar-angle maps across sessions generated by the non-selective pRF mapping stimulus on the inflated cortical surface of individuals. Standard criteria were used to identify the borders between V1, V2 & V3 (Deyoe et al., 1996; Engel et al., 1997; Sereno et al., 1995).

## Data Availability

No data is currently made available.

## Acknowledgments / Funding

We thank Matteo Lisi for valuable input into data analyses and Andrew Stockman for support in stimulus development. The research was supported by grants from the National Institute for Health Research (NIHR) Biomedical Research Centre (BRC) at Moorfields Eye Hospital NHS Foundation Trust and UCL Institute of Ophthalmology, the European Social Research Council (#ES/N000838/1); MeiraGTx, Retina UK, Moorfields Eye Hospital Special Trustees, Moorfields Eye Charity (R160035A, R190029A, R180004A), Foundation Fighting Blindness (USA) and The Wellcome Trust (099173/Z/12/Z, 100227), Ardalan Family Scholarship, the Persia Educational Foundation Maryam Mirzakhani Scholarship; and the Sir Richard Stapley Educational Trust (#313812). The views expressed are those of the authors and not necessarily those of the NHS, the NIHR, or the Department of Health.

## Author Contributions

TD and MF developed the study, collected, analysed data, and, with EA, wrote the report. EA, SS, GR, co-developed, ran and analysed a parallel study (in prep), MM initiated and co-developed the research, and with NH and SZ, identified and genotyped patients. EA, SS, AR, JG, PJ, contributed to methods/analysis development. All authors reviewed the manuscript.

## Data Sharing Statement

For GDPR and confidentiality reasons we are not able to make imaging data from paediatric participants publicly available.

## Code Availability Statement

All stimulus presentation and data analysis code are available on request to the corresponding author.

