## Supplementary Materials for "A demonstration of cone function plasticity after gene therapy in achromatopsia"

### Supplementary Materials 1: data from 2 additional treated patients

#### Participants

Here we present additional interim longitudinal measures from 2 paediatric patients with ACHM undergoing gene therapy, that replicate the findings presented in the main text. Patients T3 and T4 with CNGA3 variants, were both 10+ years old at the time of first visit (exact ages are kept undisclosed for confidentiality). T3 and T4 were tested twice, once before, and once 12 months after treatment. Pre-treatment baseline measures of patient T3 and T4 are also included in Main Figure 1 (T3=B7, T4=B6).

#### Behavioural psychophysics

Figure S1a displays binocular pre- and post-treatment thresholds of patients T3 and T4, on the 4-AFC task reported in Figure 1A&C of the main manuscript, as well as monocular post-treatment measures of patient T4. The shaded grey area indicates the 95% range of detection thresholds of the 13 untreated patients also displayed in Figure 1C. For details of the experimental procedures, see Main Methods.

Discrimination thresholds of patient T3 and T4 both were well below the upper bound of the 95% interval before treatment, with slightly better performance for patient T3. However, after treatment, Patient T4’s discrimination threshold had improved substantially to ceiling level, while patient T3’s performance had not changed measurably. Importantly, the markedly improved sensitivity to cone contrast in patient T4, tested binocularly after treatment, was driven by the treated eye; performance in the untreated eye remained at pre-treatment level. This result replicates the pattern observed in patients T1 and T2. However, whilst patient T2's discrimination threshold was amongst the highest before treatment, patient T4's was not. This shows that high pre-treatment performance on this behavioural psychophysics task is no pre-requisite for functional benefit of gene therapy.

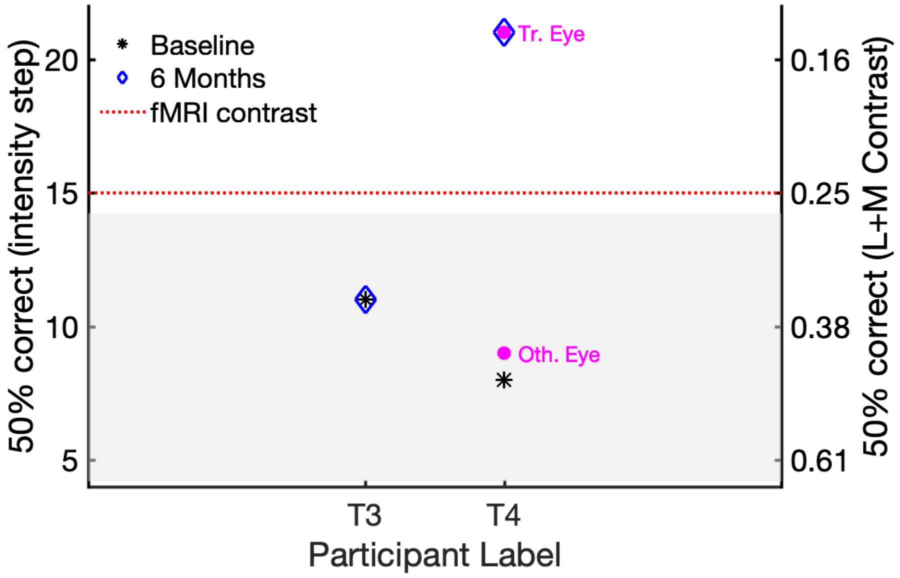


*Figure S1a. Psychophysics outside the scanner. Binocular 4AFC contrast discrimination thresholds (50% correct). Left y-axis indicates contrast detection thresholds in units of staircase step with decreasing stimulus intensity (1=highest contrast, 21=lowest contrast), with the right y-axis indicating the corresponding L+M cone Michelson contrasts (see Supplementary Material 2 for details). Stars indicate baseline measures before treatment. Shaded area: 95% prediction interval (see the main text for further details). Follow up measures ~6 months after treatment, are shown for treated case study patients T3 and T4 (blue diamonds). 6-month measures of patient T4 were repeated monocularly for the treated and other, untreated eye (magenta circles).*

**Retinotopic mapping fMRI**

After quantifying individual differences in cone function psychophysically, we tested for concurrent evidence of cone-mediated signal processing in visual cortex. As can be seen in Figure S1b, both patients showed clearly visible retinotopic organization in the rod-mediated maps (unthresholded, pRF fit $R^2 \geq 0$), with polar angle and eccentricity maps that closely resembled those 25 normal sighted control participants (Supplementary Materials 4), including the participant shown in the Main Results. Before treatment, cone-selective pRF mapping evoked no visible retinotopic map in either patient with ACHM, as expected, and in line with measures of all 11 other tested untreated patients. After treatment, patient T3 still showed no discernible cone-mediated maps, but patient T4 now demonstrated clear visual field representations in expected cortex locations, spatially aligned with the visual field representations in the rod-mediated map (Figure S1bB).

To quantify polar angle correspondence between maps, we plotted values from visual areas V1-V3 from the cone-mediated map against those from the rod-mediated map (Figure S1bC). Close correspondence between maps in polar angle layout, is reflected in the clustering of data along the identity line. To test for presence versus absence of correspondence, we compared two orthogonal regression models, one following the identity line (slope $\beta$=1, intercept=0) and the other a horizontal line with a free parameter intercept to model relationship absence. To compute the strength of evidence for the correspondence model, we calculated the Akaike Weight (AIKW) (Burnham, & Anderson, 2002; Wagenmakers et al., 2004).

Before treatment, there was poor spatial correspondence between cone- and rod-mediated polar angle maps for both patients with ACHM (T3&T4 $AICW_{\beta=1} \approx 0$), with low correlations ($CorrCoef_{Fisher\text{-}Lee}$ T3= -0.02, p<0.01, $CorrCoef_{Fisher\text{-}Lee}$ T4 = -0.01, p<0.01) and flat regression slopes ($\beta_{orth,95CI}$ T3 = [0.04:0.05]; $\beta_{orth,95CI}$ T4 = [-0.01:0.00]). ~12 months after treatment, patient T3's cone-mediated map still showed little correspondence with the retinotopic rod-mediated map ($CorrCoef_{Fisher\text{-}Lee}$ = 0.02, $\beta_{orth,95CI}$ = [-0.02:-0.01], $AICW_{\beta=1} \approx 0$). In patient T4, however, a correspondence between these maps had emerged after therapy ($CorrCoef_{Fisher\text{-}Lee}$ = 0.43, p<0.01, $\beta_{orth,95CI}$ =[ 0.76:0.78], $AICW_{\beta=1} \approx 1$) just as in patient T2.

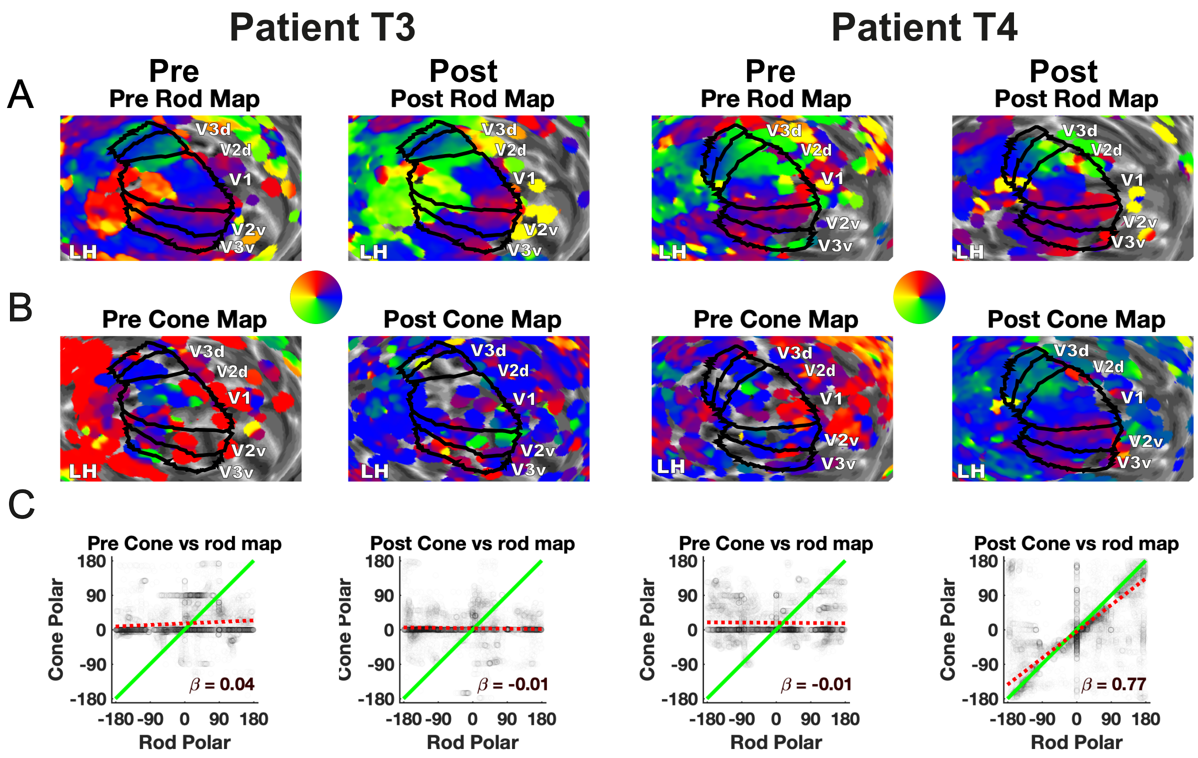


*Figure S1b. Unthresholded cone-mediated versus rod-mediated polar angle map organisation in areas V1, V2, and V3, before and after gene therapy in 2 patients. A) Rod-mediated polar angle estimates are projected onto the left hemisphere cortical surface, inflated to a sphere and zoomed in on. V1, V2, and V3 labels are drawn based on individual polar maps obtained in luminance contrast pRF scans from both sessions. B) Cone-mediated polar angle map projected on the same hemisphere C) Rod-mediated polar angle values (x-axis) from the left and right V1, V2, and V3, scattered against cone-mediated polar angle values (y-axis). Green identity line indicates perfect correspondence between the rod and cone map. Red dotted line indicates the orthogonal linear regression fit, with slope $\beta_{orth}$.*

When testing for correspondence in rod- and cone-mediated estimates of preferred eccentricity along the cortical sheet (Figure S1c), again with no statistical threshold applied ($R^2 \geq 0$), we noted emergence of eccentricity map organisation in T4's cone-mediated measures only after treatment (pre-treatment: $\beta_{orth,95CI,Ecc}$ =[0.02:0.04], $r_{Ecc}$ = 0.04, p<0.01, AICW $_{\beta=1,Ecc} \approx$ 0; post-treatment: $\beta_{orth,95CI,Ecc}$ =[0.74:0.78], $r_{Ecc}$ = 0.44, p<0.01, AICW $_{\beta=1,Ecc} \approx$ 1). However, no pattern was observed in the cone-mediated eccentricity map organisation of patient T3 before or after treatment (pre-treatment: $\beta_{orth,95CI,Ecc}$ =[-2.68:-2.40], $r_{Ecc}$ = -0.24, p<0.01, AICW $_{\beta=1,Ecc}$ = 0.19; post-treatment: $\beta_{orth,95CI,Ecc}$ =[-0.06:-0.05], $r_{Ecc}$ = -0.28, p<0.01, AICW $_{\beta=1,Ecc} \approx$ 0).

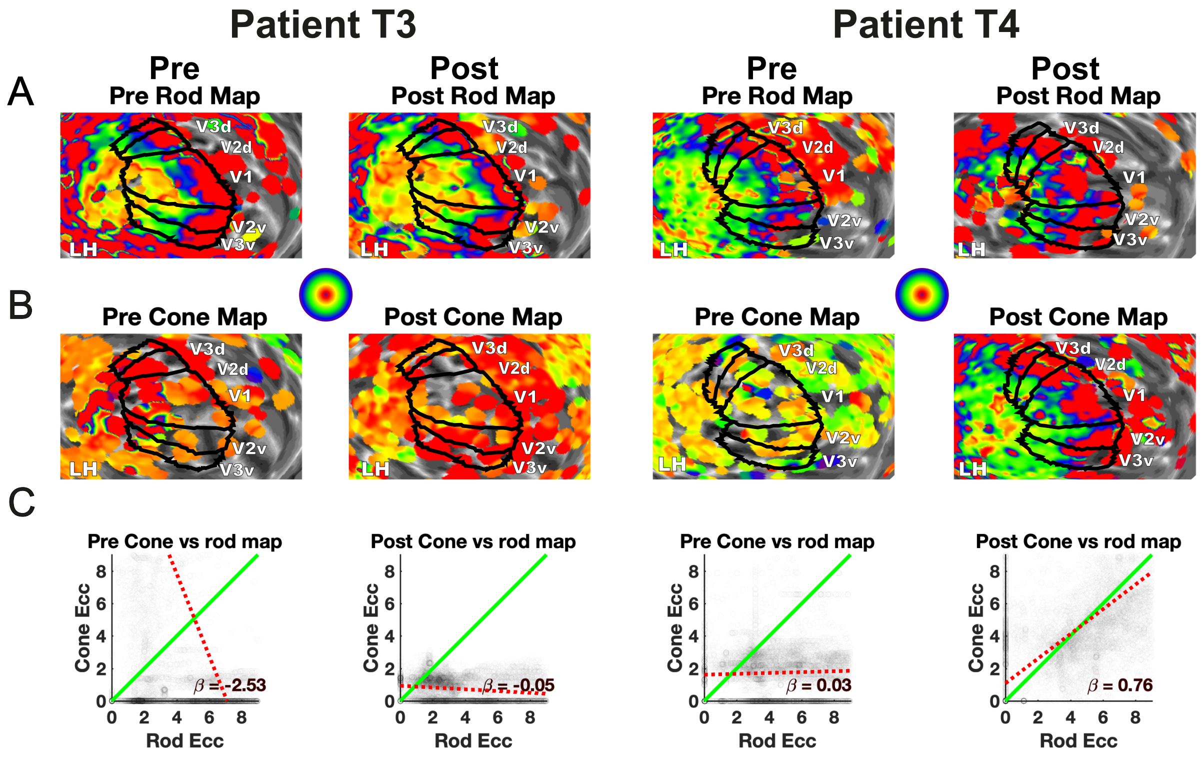


*Figure S1c. Unthresholded cone-mediated versus rod-mediated eccentricity map organisation in areas V1, V2, and V3, before and after gene therapy in 2 patients. A) Rod-mediated Eccentricity estimates are projected onto the left hemisphere cortical surface, inflated to a sphere and zoomed in on. V1, V2, and V3 labels are drawn based on individual polar maps obtained in luminance contrast pRF scans from both sessions. B) Cone-mediated eccentricity estimates projected on the same hemisphere C) Eccentricity estimates from rod-mediated map (x-axis) from left and right V1, V2, and V3, scattered against eccentricity estimates from cone-mediated map (y-axis). Green identity line indicates perfect correspondence between the rod and cone map. Red dotted line indicates the orthogonal linear regression fit, with slope* $\beta_{orth}$.

### Head movement

Head motion translations in the X, Y and Z direction are shown for the two age-matched paediatric ACHM patients, T3 and T4. Figure S1dA shows translations from cone-selective pRF mapping runs and Figure S1dB from rod-mediated runs pre- and post-treatment. Two identical fMRI runs collected at each measurement point are indicated by a solid (pRF run 1), and dashed line (pRF run 2). Small head movements and slow drifts, such as the downward drift in patient T4's pre-treatment measures, are well-accounted for using standard motion correction procedures as implemented in SPM12. Both patients also made some large head movements in the post-treatment measures. However, as these head movements were well-matched across patients and across rod- and cone-mediated scans in which maps were versus were not observed, it is highly unlikely that this factor drives the difference in cone-mediated cortical maps between patients after treatment. The relative robustness to head-movement of this measure, demonstrates usability of this approach with patients less able to stay still for extended periods.

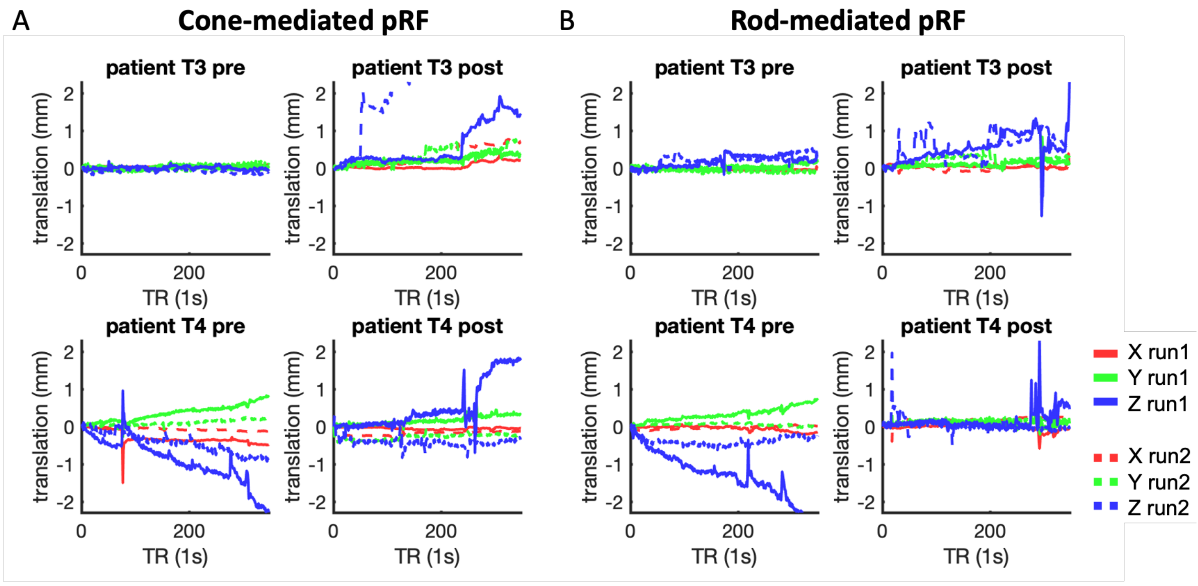


*Figure S1d. Head motion translation. Translation across X, Y, and Z direction plotted for patients T3 and T4 across rod-equated and cone-equated conditions. Each condition consisted of two runs, indicated by different line styles.*

### Eye movements

Figure S1e shows the unfiltered eye movement data of both patients T3 and T4 before and after treatment during cone-selective and rod-selective pRF mapping in the scanner. For details on how these measures were obtained from patients with nystagmus, please see Main Methods and Supplementary Materials 7. Fixation compliance was high throughout the runs, in line with high fixation task performance (>95%), and fixation observed on a face camera. It is important to note that there are substantial challenges of accurately measuring eye-movement in the MRI scanner in patients with nystagmus. Therefore, while eye movement data in patients T3 and T4 was more variable than patients T1 and T2 (presented in the main text), it is unclear whether this is due to measurement artefact (i.e., difficulty tracking the pupil, scanner vibration, drift, errors in post-hoc calibration) or substantially greater nystagmus eye movement. Crucially, variability of eye-movement measures was similar in rod-mediated scans that yielded clear maps and cone-mediated scans that yielded no maps. This is also reflected in median standard deviations of horizontal eye movements across 1-second intervals after blink and velocity outlier removal in Table S1. Based on this we conclude that the differences in cone-mediated map structure in the cortex of patient T3 and T4 before and after treatment is highly unlikely to be driven by different fixation stabilities. Moreover, given the concurrent change in psychophysics, which unlike fMRI does not require good fixation, the differences between fMRI and psychophysics measures in patients T3 and T4, likely reflect a change in cone function.

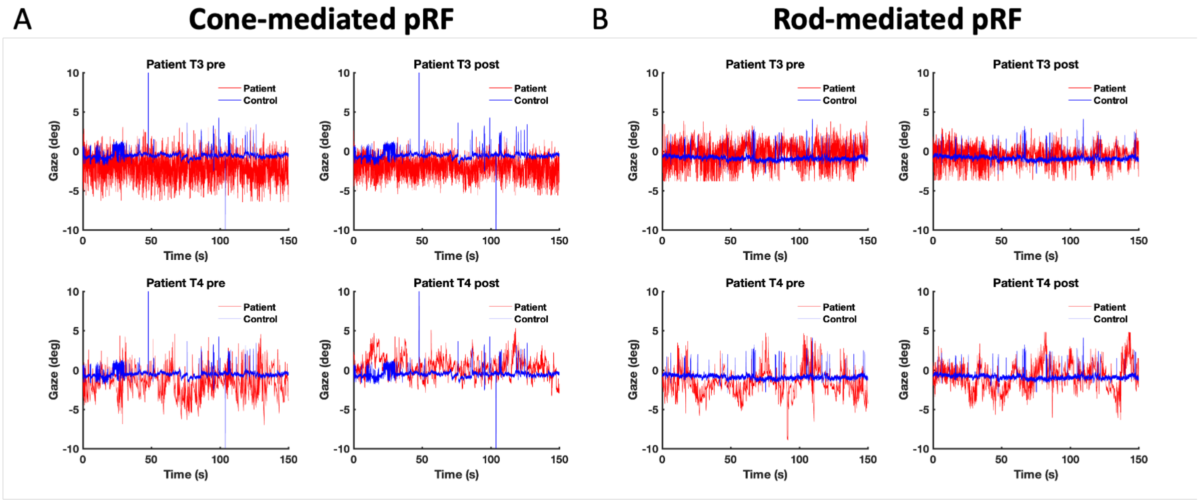


*Figure S1e. Unfiltered horizontal gaze measures across the first 150 seconds of scanning (out of 340 seconds) during the 1st data acquisition run for the: A) cone map (1st out of 2) and B) rod map (1st out of 2) in patients T3 and T4 compared to a control participant.*

| | **T3, pre** | **T3, post** | **T4, pre** | **T4, post** |
|---|---|---|---|---|
| **Rod Stim** | 0.93 | 0.69 | 0.84 | 0.75 |
| **Cone Stim** | 1.25 | 0.99 | 0.84 | 0.63 |

*Table S1: Median standard deviations of horizontal eye movement across 1-second sections along the 340-second run, after blink- and velocity outlier removal (±2SD).*

### Supplementary Materials 2: Validating cone selective stimuli

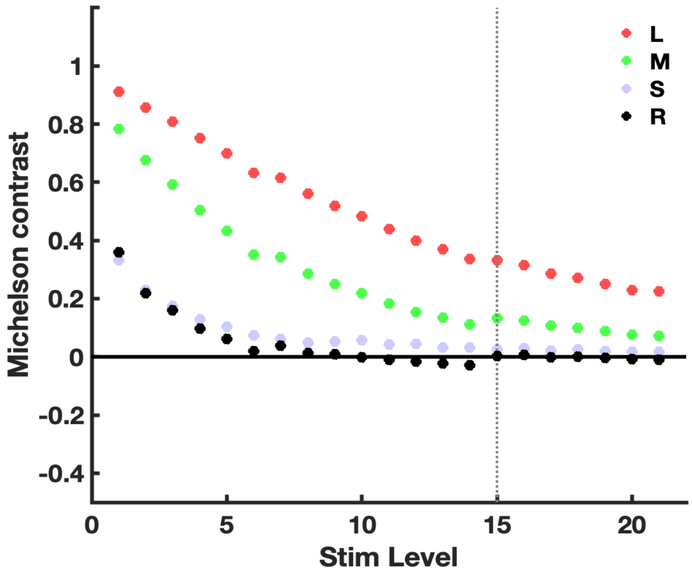


*Figure S2a. Cone selective stimuli: Michelson contrast for L-, M-, S-cones and rods, across the incrementing and decrementing RGB triplet of a chromaticity pair designed to selectively activate L and M cones. Contrasts are shown for 21 stimulus intensities designed to gradually decrease L + M contrast. Black solid line indicates zero contrast. Black dotted line indicates the contrast of the stimulus presented in the scanner.*

Cone selective chromatic pairs (shown in Main Figure 1A, B) were designed to increase (foreground) and decrease (background) cone contrast in 20 steps (see Main Methods). To quantify photoreceptor-specific contrast induced by each pair, we measured the screen spectral output for each RGB triplet used to increment/decrement the cone response, using a Spectrascan Spectroradiometer (PR-655, PhotoResearch Inc.). Each of the chromatic spectra were then multiplied with the standard observer photoreceptor fundamentals for long (L)-, mid (M)-, short (S) cone and rod (R) photoreceptors (Binder, 2008; Stockman & Sharpe, 2000) resulting in 2 activation levels (background vs. foreground) per chromatic pair for each photoreceptor type. To quantify photoreceptor-specific contrast between the foreground and background of each chromatic pair, we computed the Michelson contrast, with values near zero indicating minimal photoreceptor activation (*i.e.,* ‘silence’). Figure S2a displays the L, M, S, and R Michelson contrast across 21 chromatic pairs, designed to gradually decrease L and M cone contrast, whilst keeping rod contrast zero. As intended, the computed contrast for L-cones and M-cones decreased gradually across the stimulus range. S-cone contrast (left uncontrolled) was substantially lower, and also decreased across chromatic pairs. Figure S2a also reveals imperfect silencing of rod photoreceptors for higher stimulus contrast levels, likely due to technical issues related to imperfect correction for the non-canonical gamma function applied by the MR-compatible LCD display. Therefore, patients without functioning cones were a-priori expected to perceive the higher-contrast stimulus levels using rod-based vision. In addition, this approach rests on the assumptions that photoreceptor sensitivity functions are identical across ACHM patients, normal sighted controls, and retinal eccentricities, and that light measurements are precise and accurate. In reality, variations in photoreceptor sensitivity and measurement error, may induce deviations from the photoreceptor activation levels shown in Figure S2a, and thus limit our ability to control and verify individual photoreceptor activation levels.

Given these considerations, it is crucial that we were able to establish that chromatic pairs with lower L + M contrasts, including the contrast shown in the fMRI scanner (Level 15, shown as the vertical dotted line in Figure S2a and as the horizontal dotted red line in Figure 1C&D), were well below the detection threshold for all the untreated patients with ACHM, across the two psychophysical tasks. These individual threshold estimates had good correspondence across the two tasks (Figure S2b), even though the chromaticity pairs were embedded in spatiotemporally different stimuli and different tasks (locating a target versus discriminating checkerboard movement, Main Figure 1). Data collected in piloting and a subset of patients, also showed that measures were repeatable across staircase starting points (Figure S2c). In contrast, for normal sighted individuals, all contrasts included in the cone-selective stimulus range were above threshold and clearly visible, with performance at ceiling (level indicated by yellow square Figure S2b).

Together these measures show that the range of cone-selective stimuli used in our psychophysical tests and pRF mapping were correctly calibrated to stimulate L and M cones but not rods. We therefore conclude that improvements in contrast sensitivity beyond the pre-treatment range of performance, provides strong evidence for emerging cone function in treated ACHM.

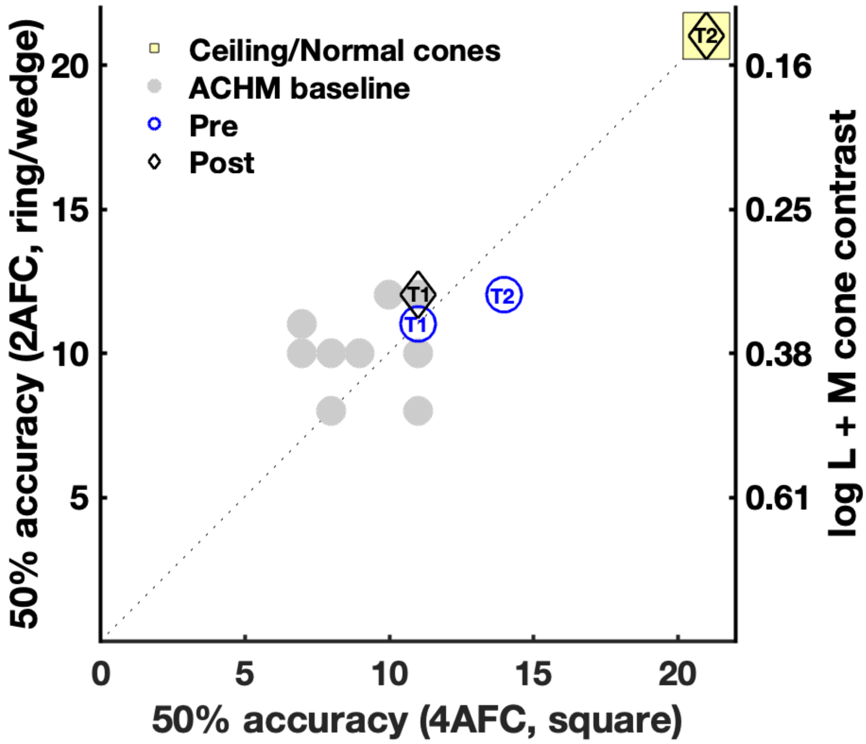


*Figure S2b: Two 50% accuracy cone sensitivity thresholds were obtained for 10 patients with ACHM using the range of L&M contrasts shown in Figure S2a. These were assessed using two different tasks: a 4AFC task that involved locating a target square (see Figure 1, main text) and a 2AFC task that involved judging movement direction of a ring-and-wedge stimulus. Thresholds on each measure for untreated baseline patients in Main Figure 1 (n=10) are scattered against each-other in grey, to visualise the test-retest range across both tasks. Notably, there was good correspondence across threshold estimates despite the different tasks and chance-performance levels for 2-AFC and 4-AFC (mean threshold difference = 0.45, maximum difference, 4 steps). Patient T2's post-intervention measurement (black diamonds) falls well outside this two-dimensional range, and at the level reached by normal sighted controls.*

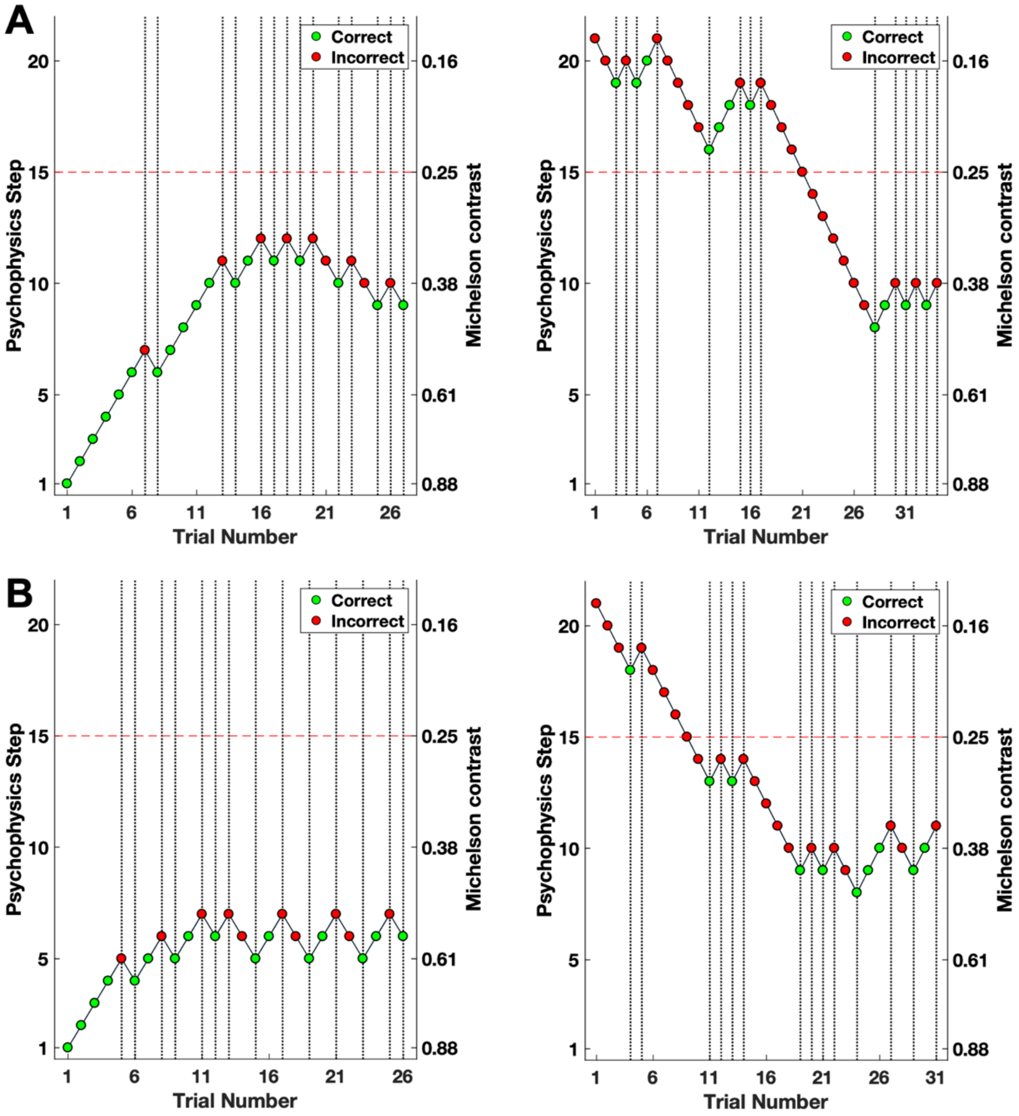


*Figure S2c. Data from two 4-AFC staircases from two participants with untreated ACHM, one with the starting point at the highest (left, step 1) and one at the lowest (right, step 21) L+M cone contrast. Above-threshold discrimination is expected for higher Michelson contrast levels (close to step 1) due to predicted rod activation for these stimuli (Figure S2a). For the participant in the lower panel (B), there is some test-retest discrepancy in this threshold (step 6 vs. 9), which may reflect measurement error, or local minima in individual rod activation. 50% accuracy thresholds remain well below the contrast level presented in the scanner (step 15), independent of staircase starting point.*

### Supplementary Materials 3: Validating rod selective stimuli

To match the rod-mediated retinotopic map as closely as possible across individuals with and without cone function, we generated RGB triplets that increased and decreased rod activation, whilst keeping L and M cone activation minimal (See Figure S3A; Estévez & Spekreijse, 1982; Spitschan & Woelders, 2018). When using three colour channels (R, G, B) only three photoreceptors types (here two cone types and rods) can be silenced simultaneously, so there was substantial S-cone contrast for the rod-activating chromatic pairs. In addition, there was some unintended L and M cone contrast, likely due to light measurement- and gamma correction error. To mitigate these unintended cone signals, we presented stimuli at the darkest light level possible (0.02 cd/m$^2$, low mesopic) to favour rod responses and render cones insensitive. The rod-mediated eccentricity map obtained with these stimuli, averaged across 26 normal-sighted control participants (all cortical surface-based aligned to the fsaverage), revealed disproportionate signal loss around the fovea (Figure S3B, left) compared to the cone-selective map in these same individuals. This suggests that we successfully mapped the central rod scotoma, by minimising, if not eliminating, responses from cone photoreceptors, which primarily populate the fovea (Figure S3B, right).

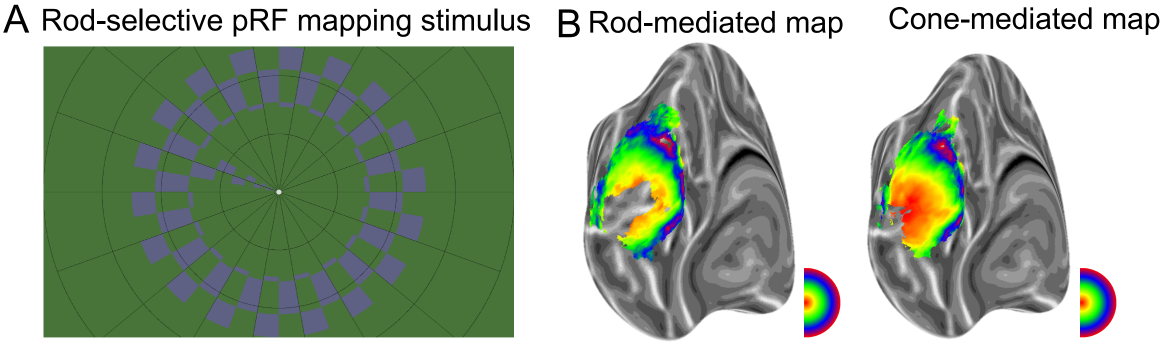


*Figure S3. A) Rod-selective pRF mapping stimulus shown in the MRI scanner. Participants detected target dimming events at fixation. B) Left: rod-mediated pRF map, obtained with stimulus in (A) averaged across 26 normally sighted control participants. Right: Cone-mediated pRF map obtained with stimulus in Main Figure 1C, averaged across 28 normally sighted control participants. Significant loss of signal around the fovea in the rod-mediated map compared to the cone-mediated map (areas coded red), suggests cone responses around the fovea were successfully reduced by the rod-selective stimulus.*

### Supplementary Materials 4: Retinotopic organisation of cone-mediated map

In Main Figure 2, we presented the polar angle estimates derived from cone-mediated pRF maps, plotted against polar angle estimates from the rod-mediated retinotopic map for patient T1, T2, and one age-matched normal sighted control. Below (left grid) we plot the same data for the additional 25 normal-sighted controls, and from 11 untreated patients with ACHM tested in addition to patients T1 and T2. To maximise the chance of detecting correspondence between the cone-mediated and rod-mediated pRFs in patients with ACHM, no threshold was applied to the pRF model goodness-of-fit ($R^2 = 0$). Reported beta values indicate the slope of an orthogonal linear regression model fit to the data (yellow dotted lines). In all normal sighted participants, data closely follow the identity line (red), indicating good alignment of retinotopic organisation between the cone- and rod-mediated maps. In contrast, data from untreated patients with ACHM show no clustering along the identity line, reflecting poor spatial correspondence of cone and rod-mediated maps.

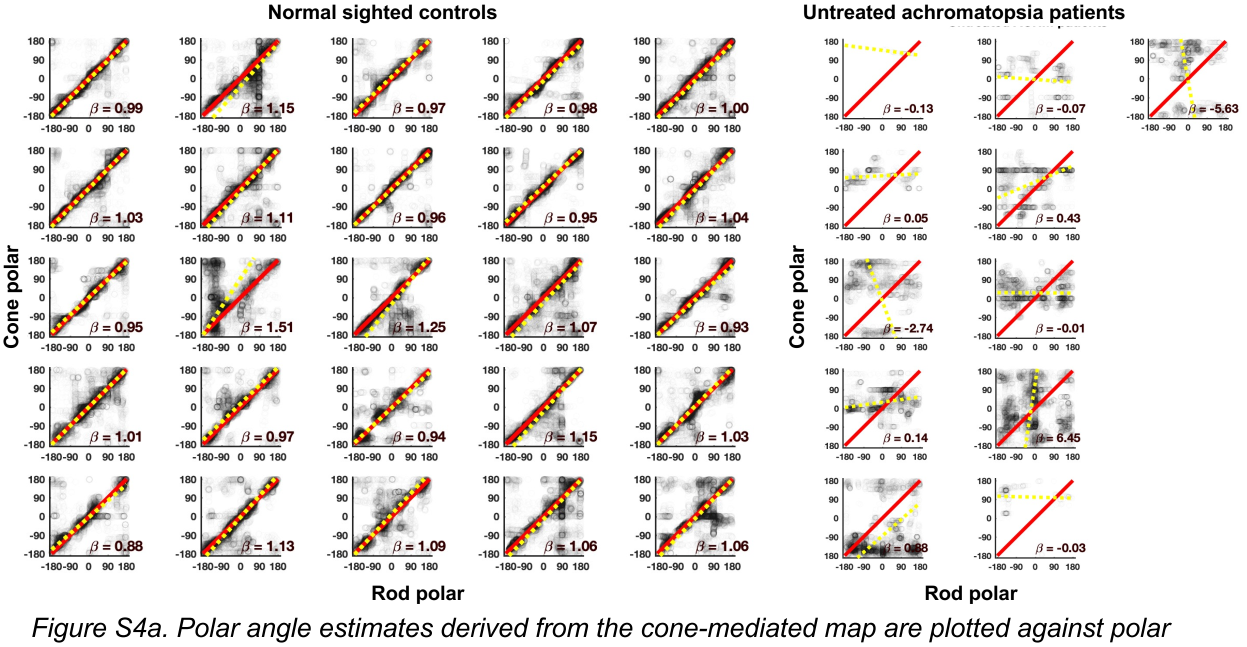


*Figure S4a. Polar angle estimates derived from the cone-mediated map are plotted against polar angle estimates from the rod-mediated retinotopic map, for normal sighted control participants (left) and untreated patients with ACHM (right). Red line indicates perfect polar angle correspondence across maps, yellow dotted line indicates best orthogonally fitting trendline.*

To verify that the differences in correspondence in polar angle values between patients with ACHM and normal sighted controls in Main Figure 2 are not an artefact of the circularity of polar angle data, we repeated this analysis on separate best-fitting X and Y coordinates of the pRF model, estimated based on the cone-selective versus rod-selective stimuli. Again, data was not corrected for goodness of fit to maximise the chance of detecting subtle retinotopic structure ($R^2 = 0$). Data in Figure S4b reveals good correspondence across the rod- and cone-mediated map of the control participant, as reflected in a high Pearson's correlation, an orthogonal linear regression slope close to 1 for both the X and Y coordinate, and an Akaike Weight (AICW; Burnham, & Anderson, 2002; Wagenmakers et al., 2004) providing strong evidence for a model with good pRF parameter correspondence versus no relationship ($\beta_{orth,X,95CI}$ = [0.95:0.97], $r_x = 0.84$, $p<0.001$, $AICW_{\beta=1,X} \approx 1$; $\beta_{orth,Y,95CI}$ = [1.26:1.30], $r_y = 0.80$, $p<0.001$, $AICW_{\beta=1,Y} \approx 1$). In contrast, there was no evidence for correspondence across the maps of untreated patient T1 ($\beta_{orth,X,95CI}$ = [-0.02:-0.005], $r_x = -0.06$, $p<0.001$, $AICW_{\beta=1,X} \approx 0$; $\beta_{orth,Y,95CI}$ = [0.02:0.03], $r_y = 0.13$, $p<0.001$; $AICW_{\beta=1,Y} \approx 0$) and patient T2 ($\beta_{orth,X0,95CI}$ = [-0.005:0.005], $r_x = -0.001$, $p=0.92$, $AICW_{\beta=1,X} \approx 0$; $\beta_{orth,Y0,95CI}$ = [0.03:0.04], $r_y = 0.14$, $p<0.001$, $AICW_{\beta=1,Y} \approx 0$). In patient T2, however, a correspondence in pRF position estimates emerged after treatment for both the X and Y coordinate, indicated by increased clustering of data along the identity line ($\beta_{orth,X,95CI}$ = [1.23:1.28], $r_{x,} = 0.66$, $p<0.001$, $AICW_{\beta=1,X\ 1} \approx 1$; $\beta_{orth,Y,95CI}$ = [1.66:1.82], $r_y = 0.34$, $p<0.001$, $AICW_{\beta=1,Y} \approx 1$). In patient T1 there was no such emergence of correspondence after treatment ($\beta_{orth,X,95CI}$ = [0.03:0.04], $r_{x,} = 0.14$, $p<0.001$, $AICW_{\beta=1,X} \approx 0$; $\beta_{orth,Y,95CI}$ = [-0.02:-0.01], $r_y = -0.03$, $p=0.02$, $AICW_{\beta=1,Y} \approx 0$). In other words, evidence for new retinotopic cone-mediated responses after treatment was present in patient T2, but not T1, confirming the results from polar angle analyses (Main Figure 2).

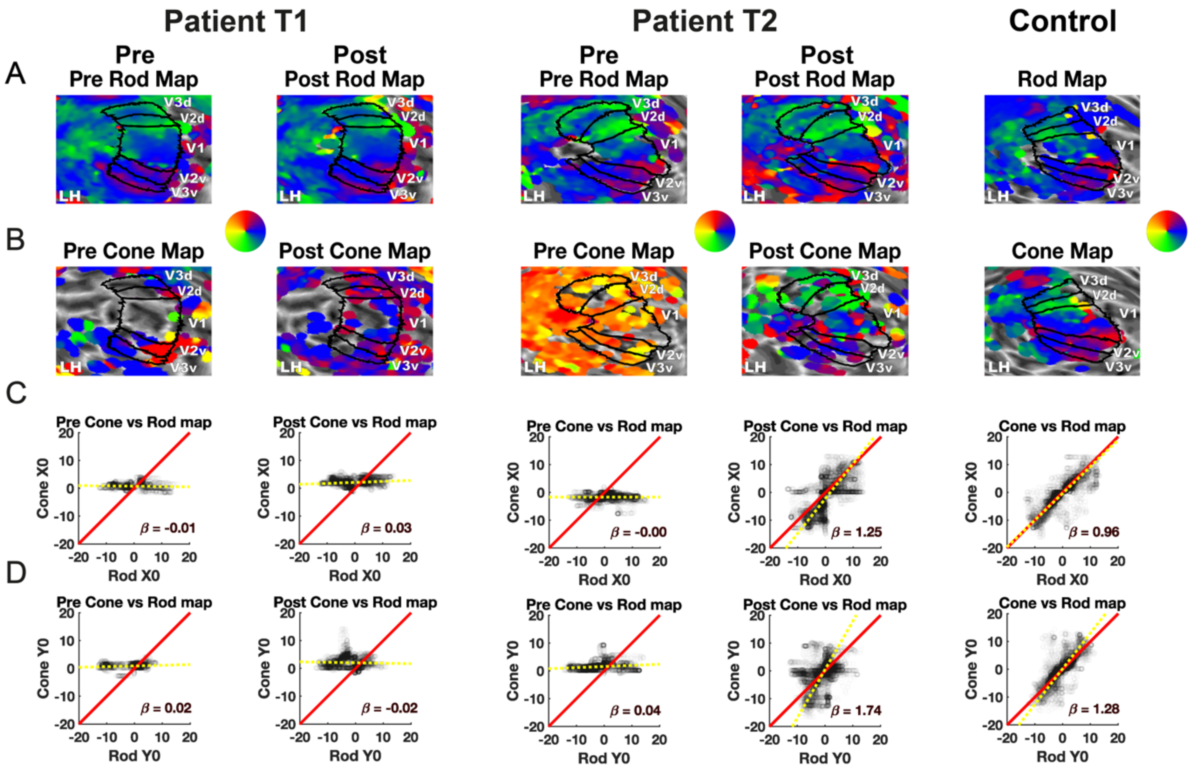


*Figure S4b. Cone-mediated versus rod-mediated X and Y pRF coordinates in degree visual angle in V1, V2, V3, before and after gene therapy in 2 patients, and an age-matched control participant. A) Rod-mediated polar angle estimates are projected onto the left hemisphere cortical surface, inflated to a sphere and zoomed in on.* V1, V2, and V3 labels are drawn based on individual polar maps obtained in luminance contrast pRF scans. *B) Cone-mediated X and Y pRF estimates projected on the same hemisphere C) X pRF coordinates from the rod-mediated map (x-axis) from left and right V1, V2, and V3, scattered against X pRF coordinates from the cone-mediated map (y-axis). Red identity line indicates perfect correspondence between the rod- and cone-mediated maps. Yellow dotted line indicates the orthogonal linear regression fit, with slope $\beta$. C) Same as B, but for Y pRF coordinates.*

When testing for correspondence in rod- and cone-mediated estimates of preferred eccentricity (Figure S4c), again with no statistical threshold ($R^2$=0), we noted a change in T2's cone-mediated measures after treatment. (pre-treatment: $\beta_{orth,95CI,Ecc}$ = -0.01:0.001, $r_{Ecc}$ = -0.09, p=0.16, $AICW_{\beta=1,Ecc} \approx 0$; post-treatment: $\beta_{orth,95CI,Ecc}$ = 3.3:4.2, $r_{Ecc}$ = 0.12, p<0.001, $AICW_{\beta=1,Ecc} \approx 1$). However, while cone-mediated eccentricity estimates in V1-V3 no longer mainly fell at ~2° after treatment, the new estimates clustered poorly around the identity line, revealing a less clear-cut correspondence between rod- and cone-mediated maps. Correspondence across the eccentricity dimension of the retinotopic map may be more sensitive to noise, or more resistant to recovery of normal organisation after life-long loss of cone function.

More data from these and other patients undergoing treatment is needed to investigate this further.

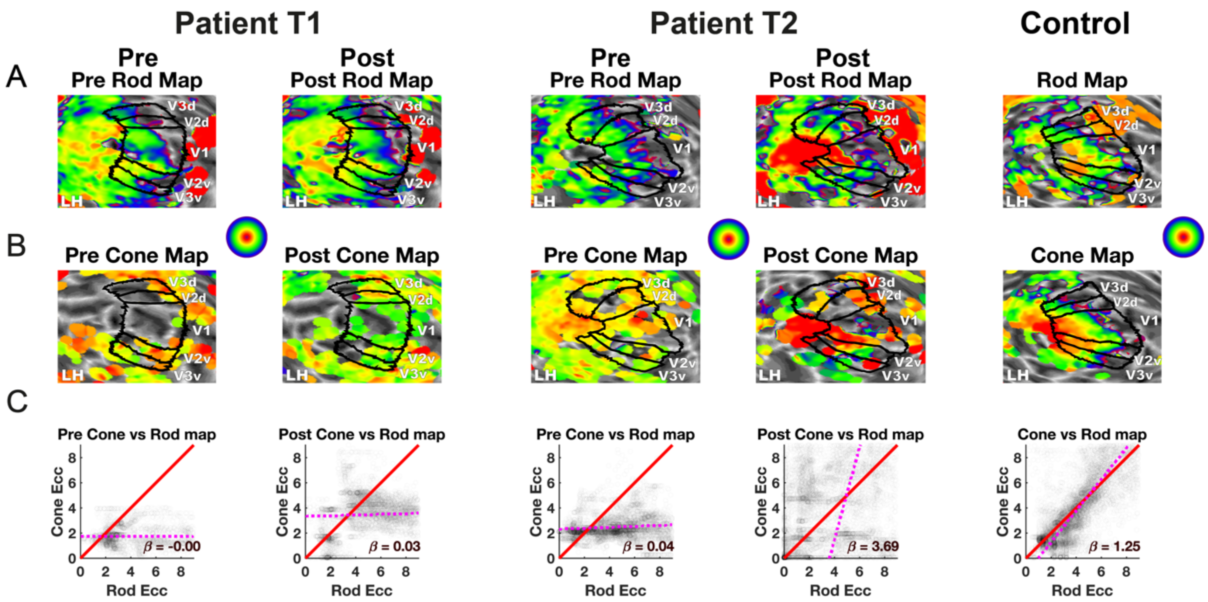


*Figure S4c. Cone-mediated versus rod-mediated eccentricity maps V1, V2, V3, before and after gene therapy in 2 patients, and an age-matched control participant. A) Rod-mediated Eccentricity estimates are projected onto the left hemisphere cortical surface, inflated to a sphere and zoomed in on.* V1, V2, and V3 labels are drawn based on individual polar maps obtained in luminance contrast pRF scans. *B) Cone-mediated eccentricity estimates projected on the same hemisphere C) Eccentricity estimates from rod-mediated map (x-axis) from left and right V1, V2, and V3, scattered against eccentricity estimates from cone-mediated map (y-axis).*

### Supplementary Materials 5: pRF size

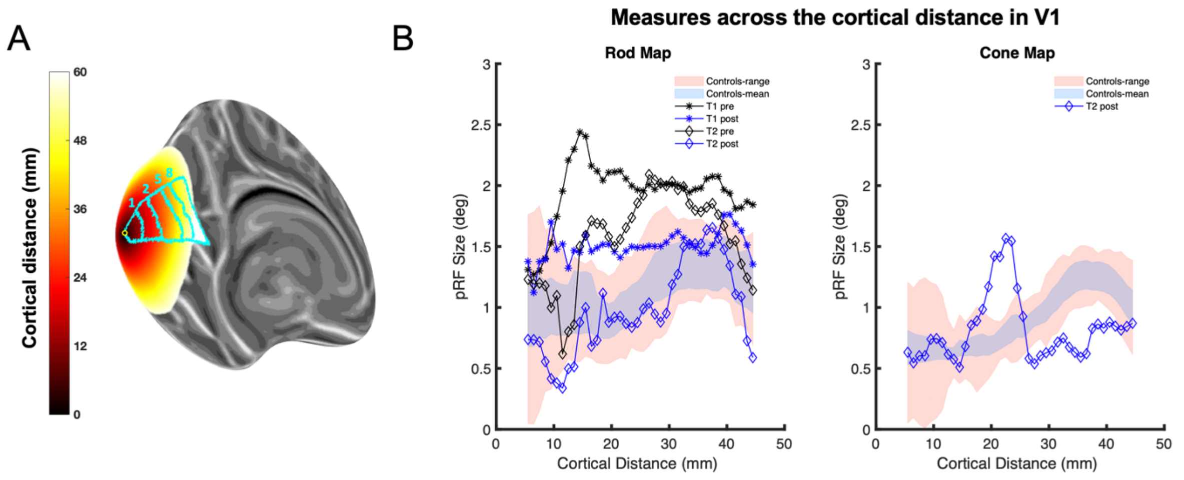


*Figure S5. A) Cortical distance across inflated left cortical hemisphere from the near-foveal origin (indicated by small circle). Eccentricity borders for 1˚, 2˚, 5˚, and 8˚ eccentricity are delineated in a label spanning from the V1 upper to lower vertical meridian bound. B) achromatopsia patients T1 (black) and T2's (blue) pRF sizes in V1, for the rod- and cone-mediated map pRFs, plotted against cortical distance from the foveal confluence. pRF size estimates for patients are referenced against a comparison measure for normal cone function (shaded pink area representing the 95% prediction interval and shaded blue area representing the 95% bootstrapped confidence interval around the mean pRF size estimate of normally sighted control participants). Measures are averaged across left and right hemispheres.*

Here, we compare the size of pRFs ($\sigma$, in degrees eccentricity) measured in area V1 from patients T1 (stars) and T2 (diamonds), before and after treatment (black and blue lines respectively), for the rod-mediated and cone-mediated pRF estimates. pRF size is plotted against cortical distance (mm) from an origin close to the foveal confluence. Estimates of pRF size for patients are referenced against those for healthy controls, for both the rod-mediated (n=26) and cone-mediated pRF maps (n=28). The shaded pink area represents the 95% prediction interval computed based on data from normally sighted control participants, and the shaded blue area represents the 95% confidence interval around the mean pRF size of these control individuals. Data with poor goodness of fit ($R^2 < 0.05$) were excluded in this analysis, as the measure of interest here is the pRF parameter itself rather than its correspondence with another independent measure. Data from the baseline cone-mediated map of patients T1 and T2, and the post-treatment cone-mediated map of patient T1 were excluded due to insufficient data after thresholding. Measures were averaged across left and right hemispheres.

Figure S5 illustrates that pRF size estimates in normal sighted controls are larger in the rod-mediated map compared to the cone-mediated map. This is consistent with lower resolution for rod-mediated signals compared to cone-mediated signals, but may also reflect different stimulus properties (*i.e.,* different contrast/luminance levels). pRF size estimates for both patients before treatment were larger than those for normal-sighted control participants, in line with low acuity associated with ACHM (Song et al., 2015). While patient T1 and T2 both had little nystagmus and showed stable fixation throughout the fMRI runs, (see Figure S7b and Table S7), effects of eye-movement on pRF size estimates (Clavagnier et al., 2015) might also contribute to group differences. After treatment, rod-mediated pRFs from the two patients with ACHM both appear to have reduced in size, irrespective of whether the patient showed indices of cone-mediated recovery of function, and despite well-matched fixation stability across sessions (see Figure S7b and Table S7). This in line with recent findings from McKyton et al. (2021) who also reported a reduction in pRF size in two adult patients in both the treated and untreated eye after gene therapy. After treatment, cone-mediated size estimates for patient T2 are similar to, or larger than those for controls (range 15 to 25 mm: approx. 1.5° to 4.5° degree eccentricity). It is important to note that pRF size estimates are more variable across sessions than position estimates, which are highly repeatable (van Dijk et al., 2016). More data is therefore needed to disentangle whether these changes in pRF size reflect random measurement variability, or treatment-related normalisation of cortical acuity for these stimuli.

### Supplementary Materials 6: Head movement

Head motion translations in the X, Y and Z direction are shown for the two age-matched paediatric ACHM patients (T1 and T2), and the age-matched normal sighted child presented as comparison in Main Figures 2&3. Figure S6A shows translations from cone-selective pRF mapping runs and Figure S6B from rod-mediated runs pre- and post-treatment. Two identical fMRI runs collected at each measurement point, are indicated by a solid (pRF run 1) line, and dashed line (pRF run 2). None of the participants made large scan-to-scan translations or rotations of head position at any point; a search for translations of more than 0.8mm translation or rotations of more than 2 degrees identified zero spikes across all runs. Any small head movements and slow drifts, such as the gradual downward drift visible in the control participant, were accounted for using standard motion correction procedures as implemented in SPM12. We conclude that the main findings are highly unlikely to be explained by head movement.

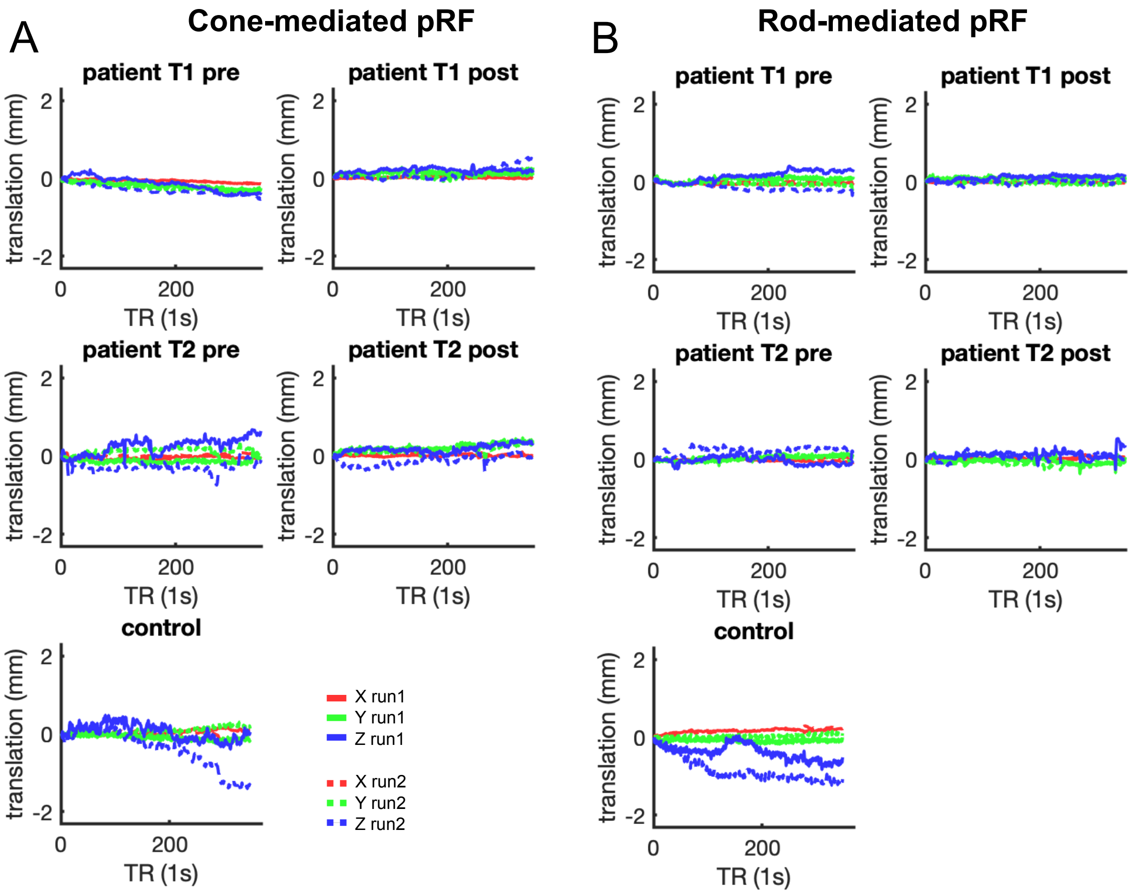


Figure S6. *Head motion translation. Translation across X, Y, and Z direction plotted for patients T1 and T2 and age-matched control participant across rod-equated and cone-equated conditions. Each condition consisted of two runs, indicated by different line styles.*

**Supplementary Materials 7: Eye movements**

Eye tracking with the Eyelink 1000 requires a built-in calibration routine, which requires high levels of fixation stability. Patients with ACHM, especially when younger, experience involuntary eye movements of varying amplitudes due to nystagmus. This makes it challenging to complete this built-in calibration, especially in children. In order to map gaze coordinates of patients in this study to screen locations during the fMRI scans, we conducted a post-hoc calibration procedure as used recently with patients with congenital nystagmus (Tailor et al., 2020), similar to other procedures (Dunn et al., 2019; Rosengren et al., 2020). We first ran a pre-calibration of the eye-tracker in the scanner using the built-in calibration on a normal sighted individual. When placing ACHM patients in the scanner, their eye was placed in a similar position as the 'pre-calibrated' eye. To account for individual differences in eye characteristics and positioning, we ran an additional in-house calibration with patients; On every other run during scanning, we asked the patients to fixate on a target presented in 5 positions across the screen in a random order (at fixation and displaced above, below, left and right, at an eccentricity of 5°). The participant was instructed to look at the fixation target for 5 seconds and move to the new location when it appeared. We then used these recordings to perform a post-hoc calibration of the data.

**Calibration:** In the post-hoc calibration, we first applied a stringent procedure to remove blinks and eye movements from the 5-point calibration recordings and identify the fixation locations for each calibration point. We first removed the first 0.5 seconds for each fixation location to allow for fixation to arrive on the target. We then removed (a) blinks and the 0.15-second period before and afterwards, when the eyelid closes and opens, (b) eye movement velocities that fell 2SD above or below the mean velocity, and (c) any positions >3SDs to the left or right of the mean fixation location, and >1SD above or below. We took the median of the remaining gaze measurements as an approximate fixation estimate. The resulting 5 median fixation locations were used to fit an affine transformation that remapped the recorded gaze positions into screen space.

**Quantifying fixation stability:** To assess fixation stability, we applied the affine transformation obtained from the post-hoc calibration to the gaze positions recorded during central fixation, whilst pRF mapping occurred in the scanner. Using data that was unfiltered for blinks and other eye movements, we first verified good compliance with the fixation task (Figure S7a). Because vertical gaze position during scanning is confounded with blink-, eye-lash- artefacts, and drift from scanner vibration, we focussed on the horizontal (x) direction, which is the dominant direction of nystagmus. Fixation compliance was high, in line with high fixation task performance (>95%), and good fixation observed on a face camera. Then, to quantify eye-movement amplitude in the horizontal direction, we removed blinks and eye movement velocity outliers (±2SD). After applying this filter, characteristic pendular nystagmus was discernible in ACHM patients, albeit with small amplitude in these two individuals with good gaze control (see Figure S7b, top panel for example). To obtain an index of fixation stability per condition and session for each participant, we computed the standard deviation of gaze position across consecutive 1-second sections of these horizontal eye position measures (see Figure S7b, bottom panel for an example), and then took the median of these measures across run 1 and 2. Results are reported in Table S7. These data revealed that both T1 and T2 had excellent control over their nystagmus in the scanner (largest median std=0.27 degrees), but that their horizontal fixation error was nevertheless about twice as large as that of the control participant.

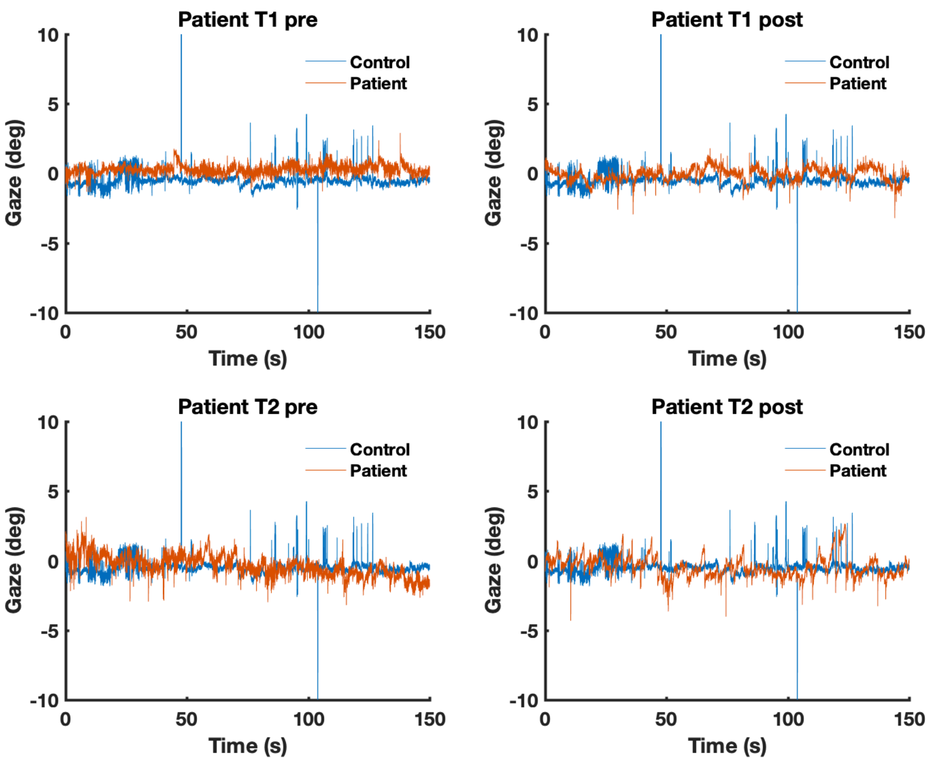


*Figure S7a: Unfiltered horizontal gaze measures across the first 150 seconds of scanning (out of 340 seconds) during the 1st data acquisition run for the cone map (1st out of 2) in patients T1 and T2 compared to a control participant.*

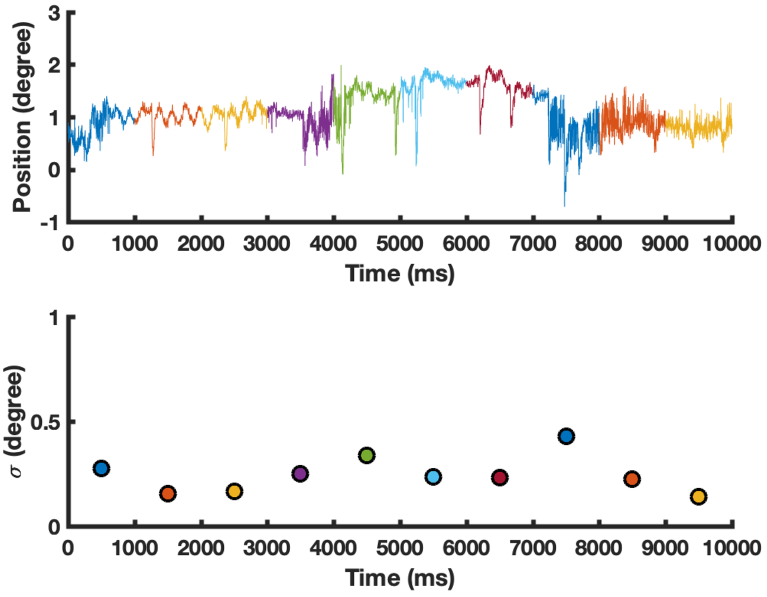


*Figure S7b: Top: Horizontal eye movement recordings from patient T1 during fMRI scanning, after the removal of blinks and recordings with velocity exceeding 2SD. Data are from first 10 seconds of fixation during the first rod-selective pRF mapping scan. 1-second consecutive sections used to compute standard deviation of gaze position in panel below are colour-coded. While a characteristic pendular movement is visible, the amplitude is very small, in line with that this participant had good*

*control over their nystagmus. Below: standard deviation computed per 1-second interval of data, across colour-coded 1-second intervals in top panel.*

| | **T1, pre** | **T1, post** | **T2, pre** | **T2, pre** | **Control** |
|---|---|---|---|---|---|
| **Rod Stim** | 0.17 | 0.27 | 0.19 | 0.25 | 0.13 |
| **Cone Stim** | 0.18 | 0.15 | 0.24 | 0.22 | 0.13 |

*Table S7: Median standard deviations of horizontal eye movement across 1-second sections along the 340-second run, after blink- and velocity outlier removal (±2SD).*

**References Supplementary Materials**


Binder, B. (2008). Color science. *Flexo*, *33*(2), 38–42. https://doi.org/10.1016/b0-12-227410-5/00945-5

Burnham, K. P. & Anderson, D. R. (2002). *Model Selection and Multimodel Inference: A Practical Information-Theoretic Approach.* Springer New York, 2002.

Clavagnier, S., Dumoulin, S. O., & Hess, R. F. (2015). Is the cortical deficit in amblyopia due to reduced cortical magnification, loss of neural resolution, or neural disorganization? *Journal of Neuroscience*, *35*(44), 14740–14755. https://doi.org/10.1523/JNEUROSCI.1101-15.2015

Dunn, M. J., Harris, C. M., Ennis, F. A., Margrain, T. H., Woodhouse, J. M., McIlreavy, L., & Erichsen, J. T. (2019). An automated segmentation approach to calibrating infantile nystagmus waveforms. *Behavior Research Methods*, *51*(5), 2074–2084. https://doi.org/10.3758/s13428-018-1178-5

Estévez, O., & Spekreijse, H. (1982). The “silent substitution” method in visual research. *Vision Research*, *22*(6), 681–691. https://doi.org/10.1016/0042-6989(82)90104-3

McKyton, A., Averbukh, E., Ohana, D. M., Levin, N., & Banin, E. (2021). Cortical Visual Mapping following Ocular Gene Augmentation Therapy for Achromatopsia. *Journal of Neuroscience*, *41*(35), 7363–7371. https://doi.org/10.1523/JNEUROSCI.3222-20.2021

Rosengren, W., Nyström, M., Hammar, B., & Stridh, M. (2020). A robust method for calibration of eye tracking data recorded during nystagmus. *Behavior Research Methods*, *52*(1), 36–50. https://doi.org/10.3758/s13428-019-01199-0

Song, C., Schwarzkopf, D. S., Kanai, R., & Rees, G. (2015). Neural population tuning links visual cortical anatomy to human visual perception. *Neuron*, *85*(3), 641–656. https://doi.org/10.1016/j.neuron.2014.12.041

Spitschan, M., & Woelders, T. (2018). The Method of Silent Substitution for Examining Melanopsin Contributions to Pupil Control. *Frontiers in Neurology*, *9*(NOV), 941. https://doi.org/10.3389/fneur.2018.00941

Stockman, A., & Sharpe, L. T. (2000). The spectral sensitivities of the middle- and long-wavelength-sensitive cones derived from measurements in observers of known genotype. *Vision Research*, *40*(13), 1711–1737.

https://doi.org/10.1016/S0042-6989(00)00021-3

Tailor, V. K., Theodorou, M., Dahlmann-Noor, A. H., & Greenwood, J. A. (2020). The effect of eye movements on visual crowding in congenital nystagmus. *Journal of Vision*, *20*(11), 711. https://doi.org/10.1167/jov.20.11.711

van Dijk, J. A., de Haas, B., Moutsiana, C., & Schwarzkopf, D. S. (2016). Intersession reliability of population receptive field estimates. *NeuroImage*, *143*, 293–303. https://doi.org/10.1016/j.neuroimage.2016.09.013

Wagenmakers, E. J., & Farrell, S. (2004). AIC model selection using Akaike weights. In *Psychonomic Bulletin and Review* (Vol. 11, Issue 1, pp. 192–196). https://doi.org/10.3758/BF03206482
